# Association between oral anticoagulants and COVID-19 related outcomes: two cohort studies

**DOI:** 10.1101/2021.04.30.21256119

**Authors:** The OpenSAFELY Collaborative, Angel YS Wong, Laurie Tomlinson, Jeremy P Brown, William Elson, Alex J Walker, Anna Schultze, Caroline E Morton, David Evans, Peter Inglesby, Brian MacKenna, Krishnan Bhaskaran, Christopher T Rentsch, Emma Powell, Elizabeth Williamson, Richard Croker, Seb Bacon, William Hulme, Chris Bates, Helen J Curtis, Amir Mehrkar, Jonathan Cockburn, Helen I McDonald, Rohini Mathur, Kevin Wing, Harriet Forbes, Rosalind M Eggo, Stephen JW Evans, Liam Smeeth, Ben Goldacre, Ian J Douglas

## Abstract

**Objectives:** We investigated the role of routinely prescribed oral anticoagulants (OACs) in COVID-19 outcomes, comparing current OAC use versus non-use in Study 1; and warfarin versus direct oral anticoagulants (DOACs) in Study 2.

**Design:** Two cohort studies, on behalf of NHS England.

**Setting:** Primary care data and pseudonymously-linked SARS-CoV-2 antigen testing data, hospital admissions, and death records from England.

**Participants:** Study 1: 70,464 people with atrial fibrillation (AF) and CHA□DS□-VASc score of 2. Study 2: 372,746 people with non-valvular AF.

**Main outcome measures:** Time to test for SARS-CoV-2, testing positive for SARS-CoV-2, COVID-19 related hospital admission, COVID-19 deaths or non-COVID-19 deaths in Cox regression.

**Results:** In Study 1, we included 52,416 current OAC users and 18,048 non-users. We observed no difference in risk of being tested for SARS-CoV-2 associated with current use (adjusted HR, 1.01, 95%CI, 0.96 to 1.05) versus non-use. We observed a lower risk of testing positive for SARS-CoV-2 (adjusted HR, 0.73, 95%CI, 0.60 to 0.90), and COVID-19 deaths (adjusted HR, 0.69, 95%CI, 0.49 to 0.97) associated with current use versus non-use. In Study 2, we included 92,339 warfarin users and 280,407 DOAC users. We observed a lower risk of COVID-19 deaths (adjusted HR, 0.79, 95%CI, 0.76 to 0.83) associated with warfarin versus DOACs. Similar associations were found for all other outcomes.

**Conclusions:** Among people with AF and a CHA□DS□-VASc score of 2, those receiving OACs had a lower risk of receiving a positive COVID-19 test and severe COVID-19 outcomes than non-users; this might be explained by a causal effect of OACs in preventing severe COVID-19 outcomes or more cautious behaviours leading to reduced infection risk. There was no evidence of a higher risk of severe COVID-19 outcomes associated with warfarin versus DOACs in people with non-valvular AF regardless of CHA□DS□-VASc score.

**Key points:** *What is already known on this topic:*

- Current studies suggest that prophylactic or therapeutic anticoagulant use, particularly low molecular weight heparin, lower the risk of pulmonary embolism and mortality during hospitalisation among patients with COVID-19.
- Reduced vitamin K status has been reported to be correlated with severity of COVID-19. This could mean that warfarin, as a vitamin K antagonist, is associated with more severe COVID-19 disease than non-vitamin K anticoagulants.

*What this study adds:*

- In 70,464 people with atrial fibrillation, at the threshold of being treated with an OAC based on risk of stroke, we observed a lower risk of testing positive for SARS-CoV-2 and COVID-19 related deaths associated with routinely prescribed OACs, relative to non-use.
- This might be explained by OACs preventing severe COVID-19 outcomes, or more cautious behaviours and environmental factors reducing the risk of SARS-CoV-2 infection in those taking OACs.
- In 372,746 people with non-valvular atrial fibrillation, there was no evidence of a higher risk of severe COVID-19 outcomes associated with warfarin compared with DOACs.

## Introduction

Early studies reported that anticoagulation, particularly low molecular weight heparin, lowers the risk of pulmonary embolism and mortality during hospitalisation among patients with COVID-19.^1–3^ While anticoagulants appear to be a useful treatment option for hospitalised patients with COVID-19, most studies investigated the potential protective role of routine use of oral anticoagulants (OACs) in COVID-19 related outcomes were of small sample size or in hospital setting only.^4–14^ Notably, people taking oral anticoagulants are likely to have more comorbidities than those not, and so comparing COVID-19 outcomes for OAC use with no use could be subject to confounding. Importantly, use of OAC treatment is partly threshold based; according to the guidelines for the management of patients with atrial fibrillation (AF), which is also one of the Quality and Outcome Framework indicators a payment incentive scheme in English general practice.^15–17^ People with a CHA_2_DS_2_-VASc score (used to predict risk of stroke) of ≥2 should be offered an anticoagulant. For those with a score of exactly 2, there is possibly a degree of variation in OAC prescribing, offering a useful group in whom OAC use can be compared with no use. A better understanding of the impact of OACs on COVID outcomes may alter the balance of benefits and risks for those around such a threshold.

In addition, reduced vitamin K status has been reported as a consequence of COVID-19 infection, with a correlation between vitamin K status and severity of COVID-19.^18^ This might have an implication for warfarin users (a vitamin K antagonist and anticoagulant); warfarin depletes functional vitamin K reserves for the synthesis of active clotting factors, and might therefore be associated with more severe COVID-19 disease. Unlike warfarin, direct oral anticoagulants (DOACs), do not act on vitamin K pathways to prevent blood clots. They might be favoured for treating coagulation related disorders whilst are also potentially associated with a lower risk of serious COVID-19 related outcomes. To date, the clinical evidence of the effects of warfarin compared with DOACs on COVID-19 related outcomes is lacking.

We therefore set out to investigate 1) the association between routine use of OACs and COVID-19 related outcomes compared with non-use among people with AF who had a CHA□DS□-VASc score of 2; 2) the association between warfarin and COVID-19 related outcomes, compared with DOACs among patients with non-valvular AF regardless of CHA□DS□-VASc score.

## Methods

### Study design

We conducted two cohort studies between 1^st^ March 2020 and 28^th^ September 2020.

### Data Source

Primary care records managed by the software provider TPP were linked to SARS-CoV-2 antigen testing data from the Second Generation Surveillance System, COVID-19 related hospital admissions from the secondary uses service, and Office for National Statistics death data through OpenSAFELY, a data analytics platform created by our team on behalf of NHS England.^19^ The dataset analysed within OpenSAFELY is based on 24 million people currently registered with primary care practices using TPP SystmOne software, representing 40% of the English population. It includes pseudonymised data such as coded diagnoses, prescribed medications and physiological parameters.

### Study Populations and exposure

#### Study 1: Risk of COVID-19 outcomes comparing OAC use with non-use

We identified people who had a diagnosis of AF on or before study start date (1^st^ March 2020). In order to reduce confounding by indication, we limited the cohort to those who had a CHA□DS□-VASc score of 2 as the indication for OAC therapy would typically be borderline among people with this score. We calculated their CHA□DS□-VASc score based on their demographics and recorded diagnoses.

People with missing data for gender, Index of Multiple Deprivation, <1 year of primary care records, or aged <18 or >110 and prescribed injectable anticoagulants 4 months before study start date were excluded.

We defined current OAC users as those ever prescribed OACs in the 4 months prior to study start, and non-users are those with no record of OAC prescription in the same time period.

#### Study 2: Risk of COVID-19 outcomes and non-COVID-19 death comparing warfarin with DOACs

We identified people with a diagnosis of AF on or before study start date. All patients with AF were included, regardless of CHA□DS□-VASc score.

The same exclusion criteria applied to Study 1 were applied. In addition, people with mitral stenosis or prosthetic mechanical valves, chronic kidney disease stage V (estimated glomerular filtration rate < 15mL/min or on dialysis), or antiphospholipid antibody syndrome were also excluded in this study because DOACs are less commonly used in these patient groups.

We defined participants as DOAC users if they were prescribed a DOAC as their latest OAC prescription in the 4 months before study start date. The comparison group was people who were prescribed warfarin as the latest OAC prescription in the 4 months before study start date. If both warfarin and DOACs were prescribed on the same day as the latest prescription (n=32), we classified them as warfarin users.

### Outcomes and follow-up

The outcomes were 1) tested for SARS-CoV-2, 2) testing positive for SARS-CoV-2, 3) COVID-19 related hospital admission, and 4) COVID-19 related death (defined as the presence of ICD-10 codes U071 (confirmed COVID-19) and U072 (suspected COVID-19) anywhere on the death certificate). Testing outcomes were obtained from the UK’s Pillar 1 (NHS and Public Health England laboratories) and Pillar 2 (commercial partners) testing strategies and included results from polymerase chain reaction (PCR) swab tests used to identify symptomatic individuals.^30,31^ In Study 2, we included non-COVID-19 death as a negative control outcome. We further conducted *post-hoc* analyses to include cause-specific deaths as outcomes (i.e. death due to myocardial infarction, ischaemic stroke, venous thromboembolism, gastrointestinal bleeding and intracranial bleeding) to aid the interpretation for the observed lower risk of non-COVID-19 death associated with warfarin versus DOACs.

In both studies, follow up for each cohort began on the 1^st^ March 2020 and ended at the latest of the outcome of interest in each analysis, deregistration from the TPP practice, death or study end date (28 September 2020) (Supplementary figure 1).

### Covariates

Covariates were pre-specified, identified from a Directed Acyclic Graph (DAG) approach (Supplementary figures 2 and 3), including age, sex, obesity, smoking status, hypertension, heart failure, myocardial infarction, peripheral arterial disease, stroke/transient ischemic attack, venous thromboembolism, diabetes, flu vaccination, current antiplatelet use, current oestrogen and oestrogen-like therapy use, Index of Multiple Deprivation, care home residence (for Study 2). We identified these covariates that are both associated with the exposure and the risk of severe COVID-19 outcomes^19^. Some covariates that are associated with the exposure and venous thromboembolism, possibly leading to severe COVID-19 outcomes were also included.^20,21^ All codelists for identifying exposures, covariates and outcomes are openly shared at https://codelists.opensafely.org/ for inspection and re-use.

### Statistical Methods

Baseline characteristics in each study were summarised using descriptive statistics, stratified by exposure status. We present adjusted cumulative incidence/mortality curves using the Royston-Parmar model. We estimated hazard ratios (HRs) with 95% confidence intervals (CIs) using Cox regression with time since cohort entry as the underlying timescale. We accounted for competing risk by modelling the cause-specific hazard (i.e. censoring other deaths for COVID-19 death analysis, and censoring any death for other outcomes analysis). We used graphical methods and tests based on Schoenfeld residuals to explore violations of the proportional hazards assumption.

We performed unadjusted models, models adjusted for age (using restricted cubic splines) and sex, DAG adjusted model, and fully-adjusted models (stratified by general practice). In Study 2 comparing clinical outcomes between warfarin and DOACs, the DAG adjusted model was also stratified by general practice.

### Quantitative Bias Analysis

We considered that OAC users and warfarin users may lower their COVID-19 risk through more risk-averse health behaviour (e.g. wearing face masks, avoiding close proximity to others) than, respectively, non-users and DOAC users. Given that health behaviour is not captured in medical records, we conducted quantitative bias analyses to assess the sensitivity of our results to this potential unmeasured confounder.

We estimated how strongly associated higher-risk health behaviour would need to be with each outcome, and with non-use and DOAC use, relative to OAC use and warfarin use respectively. We calculated the minimum strength of association required between an unmeasured confounder and one of exposure or outcome to move from the observed hazard ratio to a null bias-adjusted hazard ratio (i.e. the E-value).^22^ We also calculated the minimum strength of association required between unmeasured confounder and both of exposure and outcome to move from the observed hazard ratio to a null bias-adjusted hazard ratio (i.e. the Cornfield condition)^22^. Furthermore, we calculated the minimum strength of association required to move from the observed protective associations to a bias-adjusted harmful association of HR 1.2 for Study 2 because we hypothesised that warfarin is associated with a higher risk of COVID-19 related outcomes versus DOACs.

### Sensitivity Analyses

Table 1 shows the list of sensitivity analyses.

**Table 1:** List of sensitivity analyses

| Sensitivity analysis | Justification |
| --- | --- |
| In both cohort studies: |  |
| 1. Additionally adjusted for ethnicity in DAG and fully-adjusted models. In the fully adjusted models, additional covariates included chronic obstructive pulmonary disease, other respiratory diseases (not including asthma), cancer, immunosuppression, chronic kidney disease, General Practice attendance rate in the year prior to cohort entry, and Accident & Emergency attendance rate in the year prior to cohort entry. | In the main analysis, we did not adjust for ethnicity as a sizable proportion of individuals with missing ethnicity (~23%). We undertook complete case analysis to address missing data. |
| 2. Repeated main analysis excluding people prescribed antiplatelets 4 months before study start date | To explore the impact of use of antiplatelet which can reduce the risk of blood clots. |
| Cohort study 1: comparing risk of COVID-19 outcomes comparing OAC with non-use |  |
| 3. Limited the study cohort to people who aged 55 or above | To explore the impact of potential confounders in young age, for example, pregnancy. |
| 4. Stratified the cohort according to the care home residence for the outcome of testing positive for SARS-CoV-2 (*post-hoc* analysis) | To explore the impact of health behaviour as people living in care homes were less likely to have differences in behaviour. |
| Cohort study 2: comparing risk of COVID-19 outcomes comparing warfarin with DOACs |  |
| 5. Repeated main analysis excluding people who were prescribed both warfarin and DOACs on the day of the latest OAC prescription | To assess the sensitivity of exposure definition. |
| 6. Repeated main analysis excluding people who ever had warfarin prescription 4 months before study start date in the DOAC group | As warfarin is hypothesised to have harmful effect on severe COVID-19 compared with DOAC, this analysis was to assess the sensitivity of exposure definition. |
| 7. Time-updated the OAC exposure variable | To evaluate the impact of national recommendation on drug switching from warfarin to DOACs due to COVID-19 pandemic.<sup>38</sup> |

### Software and Reproducibility

Data management was performed using Python 3.8 and SQL, with analysis carried out using Stata 16.1. All study analyses were pre-planned unless otherwise stated. All code for data management and analyses in addition to the pre-specified protocol (https://github.com/opensafely/anticoagulants-research/blob/master/protocol/Protocol_%20Anticoag%20OpenSAFELY_v3.docx) are archived at: https://github.com/opensafely/anticoagulants-research. Deviations from pre-specified protocol, with reasons were stated in the Supplementary.

### Patient and Public Involvement

Patients were not formally involved in developing this specific study design that was developed rapidly in the context of a global health emergency. We have developed a publicly available website https://opensafely.org/ through which we invite any patient or member of the public to contact us regarding this study.

### Ethical approval

This study was approved by the Health Research Authority (REC reference 20/LO/0651) and by the LSHTM Ethics Board (reference 21863).

## Results

Supplementary figure 4 shows the flowchart of inclusion of people in both cohort studies.

### Main analysis

#### Study 1: Risk of COVID-19 outcomes comparing OAC use with non-use

Among 70,464 people with AF who had a CHA□DS□-VASc score of 2, we included 52,416 current OAC users and 18,048 non-users (Table 2). Median age was 71 years (IQR, 66 to 75) among current users and 69 years (IQR, 63 to 74) among non-users.

**Table 2:** Demographic and Clinical Characteristics in two cohort studies

|  | Study 1 |  | Study 2 |  |
| --- | --- | --- | --- | --- |
|  | Non-use of oral anticoagulants | Current use of oral anticoagulants | Current use of direct oral anticoagulant | Current use of warfarin |
| **Total** | 18,048 | 52,416 | 280,407 | 92,339 |
| **Age as of 1<sup>st</sup> Mar2020** |  |  |  |  |
| 18-<40 | 169 (0.9) | 126 (0.2) | 510 (0.2) | 112 (0.1) |
| 40-<50 | 615 (3.4) | 760 (1.4) | 2,320 (0.8) | 361 (0.4) |
| 50-<60 | 2,372 (13.1) | 4,682 (8.9) | 12,788 (4.6) | 2,245 (2.4) |
| 60-<70 | 6,134 (34.0) | 15,809 (30.2) | 42,407 (15.1) | 9,824 (10.6) |
| 70-<80 | 6,233 (34.5) | 22,735 (43.4) | 98,848 (35.3) | 34,051 (36.9) |
| 80+ | 2,525 (14.0) | 8,304 (15.8) | 123,534 (44.1) | 45,746 (49.5) |
| Median, IQR | 69 (63-74) | 71 (66-75) | 78 (71-84) | 79 (73-85) |
| **Sex** |  |  |  |  |
| Female | 5,345 (29.6) | 10,894 (20.8) | 122,778 (43.8) | 36,414 (39.4) |
| **Body mass index** |  |  |  |  |
| <18.5 | 267 (1.5) | 502 (1.0) | 5,437 (1.9) | 1,199 (1.3) |
| 18.5-24.9 | 4,701 (26.0) | 11,826 (22.6) | 72,658 (25.9) | 21,998 (23.8) |
| 25-29.9 | 5,948 (33.0) | 17,973 (34.3) | 94,621 (33.7) | 31,981 (34.6) |
| 30-34.9 | 3,277 (18.2) | 11,104 (21.2) | 57,590 (20.5) | 19,592 (21.2) |
| 35-39.9 | 1,401 (7.8) | 4,771 (9.1) | 24,032 (8.6) | 8,114 (8.8) |
| 40+ | 741 (4.1) | 2,882 (5.5) | 12,586 (4.5) | 4,539 (4.9) |
| Missing | 1,713 (9.5) | 3,358 (6.4) | 13,483 (4.8) | 4,916 (5.3) |
| **Ethnicity** |  |  |  |  |
| White | 12,926 (71.6) | 38,197 (72.9) | 201,046 (71.7) | 66,800 (72.3) |
| Mixed | 48 (0.3) | 102 (0.2) | 548 (0.2) | 115 (0.1) |
| Asian/Asian British | 331 (1.8) | 512 (1.0) | 3,911 (1.4) | 766 (0.8) |
| Black | 125 (0.7) | 183 (0.3) | 1,289 (0.5) | 258 (0.3) |
| Other | 92 (0.5) | 217 (0.4) | 1,100 (0.4) | 281 (0.3) |
| Missing | 4,526 (25.1) | 13,205 (25.2) | 72,513 (25.9) | 24,119 (26.1) |
| **Index of Multiple Deprivation** |  |  |  |  |
| 1 (least deprived) | 3,727 (20.7) | 11,355 (21.7) | 57,570 (20.5) | 17,703 (19.2) |
| 2 | 3,711 (20.6) | 11,030 (21.0) | 56,881 (20.3) | 18,400 (19.9) |
| 3 | 3,624 (20.1) | 10,530 (20.1) | 55,654 (19.8) | 19,056 (20.6) |
| 4 | 3,548 (19.7) | 9,951 (19.0) | 54,758 (19.5) | 18,615 (20.2) |
| 5 (most deprived) | 3,438 (19.0) | 9,550 (18.2) | 55,544 (19.8) | 18,565 (20.1) |
| **Smoking status** |  |  |  |  |
| Never | 6,908 (38.3) | 19,002 (36.3) | 101,492 (36.2) | 33,005 (35.7) |
| Former | 9,309 (51.6) | 29,430 (56.1) | 161,752 (57.7) | 54,463 (59.0) |
| Current | 1,780 (9.9) | 3,894 (7.4) | 16,828 (6.0) | 4,834 (5.2) |
| Missing | 51 (0.3) | 90 (0.2) | 335 (0.1) | 37 (0.0) |
| **Hazardous alcohol use** | 2,339 (13.0) | 6,501 (12.4) | 28,375 (10.1) | 7,819 (8.5) |
| **Care home residence\*** | 215 (1.2) | 325 (0.6) | 8,133 (2.9) | 1,039 (1.1) |
| **Comorbidities** |  |  |  |  |
| Hypertension | 7,500 (41.6) | 21,851 (41.7) | 195,078 (69.6) | 66,888 (72.4) |
| Heart Failure | 1,393 (7.7) | 5,624 (10.7) | 71,427 (25.5) | 26,926 (29.2) |
| Myocardial infarction | 754 (4.2) | 1,612 (3.1) | 31,911 (11.4) | 10,414 (11.3) |
| Peripheral arterial disease | 209 (1.2) | 424 (0.8) | 14,273 (5.1) | 5,091 (5.5) |
| Stroke/transient ischaemic attack | 333 (1.8) | 1,029 (2.0) | 60,271 (21.5) | 18,470 (20.0) |
| Venous thromboembolism | 318 (1.8) | 548 (1.0) | 19,927 (7.1) | 8,202 (8.9) |
| Diabetes |  |  |  |  |
| Controlled (HbA1c < 58 mmols/mol) | 1,805 (10.0) | 4,246 (8.1) | 61,178 (21.8) | 23,893 (25.9) |
| Uncontrolled (HbA1c ≥ 58 mmols/mol) | 768 (4.3) | 2,094 (4.0) | 22,672 (8.1) | 7,696 (8.3) |
| HbA1c not measured | 62 (0.3) | 72 (0.1) | 838 (0.3) | 298 (0.3) |
| COPD | 1,523 (8.4) | 5,216 (10.0) | 36,189 (12.9) | 11,272 (12.2) |
| Other respiratory diseases | 741 (4.1) | 2,110 (4.0) | 16,444 (5.9) | 4,731 (5.1) |
| Cancer | 2,779 (15.4) | 7,930 (15.1) | 49,488 (17.6) | 16,240 (17.6) |
| Immunosuppression | 257 (1.4) | 438 (0.8) | 1,688 (0.6) | 528 (0.6) |
| Chronic kidney disease | 2,031 (11.3) | 8,431 (16.1) | 95,715 (34.1) | 34,633 (37.5) |
| **Primary care consultations** |  |  |  |  |
| Median, IQR | 7 (3-12) | 10 (5-16) | 10 (6-17) | 16 (9-27) |
| Min, Max | 0, 202 | 0, 198 | 0, 432 | 0, 307 |
| **A&E attendance** |  |  |  |  |
| Median, IQR | 0 (0-0) | 0 (0-1) | 0 (0-1) | 0 (0-1) |
| Min, Max | 0, 39 | 0, 54 | 0, 69 | 0, 45 |
| **Flu vaccination** | 11,064 (61.3) | 39,613 (75.6) | 220,153 (78.5) | 78,558 (85.1) |
| **Medications** |  |  |  |  |
| Oestrogen/<br>oestrogen-like drugs | 246 (1.4) | 352 (0.7) | 1,652 (0.6) | 361 (0.4) |
| Antiplatelets | 3,951 (21.9) | 2,296 (4.4) | 19,030 (6.8) | 4,108 (4.4) |

Abbreviations: COPD, Chronic obstructive pulmonary disease; DMARDs, Disease-modifying antirheumatic drugs
\*Numbers are round to nearest 5 for the cohort in Study 1 as the count for missing value was between 1-5.

Current OAC users were more likely to be men, obese, former smokers, and have a medical history of heart failure, chronic obstructive pulmonary disease, chronic kidney disease than non-users. Current users were less likely to have a medical history of myocardial infarction, peripheral artery disease, venous thromboembolism, immunosuppression, and diabetes than non-users. Current users were also less likely to have a prescription for oestrogen or oestrogen-like drugs, antiplatelets, non-steroidal anti-inflammatory drugs, and aspirin, but to have had more primary care consultations and vaccinations than non-users.

Supplementary figure 5 presents time to COVID-19 outcomes in adjusted cumulative incidence plots. For being tested for SARS-CoV-2, the unadjusted HR for current use of OACs was 0.96 (95% CI, 0.92 to 1.00), with a DAG-adjusted HR of 1.01 (95% CI, 0.96 to 1.05), versus non-use (Figure 1 & Supplementary table 2). A lower risk of testing positive for SARS-CoV-2 [unadjusted HR: 0.75 (95% CI, 0.62 to 0.92); DAG adjusted HR: 0.73 (95% CI, 0.60 to 0.90)], COVID-19 related hospital admission with wide confidence interval [unadjusted HR: 0.93 (95% CI, 0.69 to 1.24); DAG adjusted HR: 0.86 (95% CI, 0.63 to 1.17)], and COVID-19 related deaths [unadjusted HR: 0.81 (95% CI, 0.59 to 1.11); DAG adjusted HR: 0.69 (95% CI, 0.49 to 0.97)], comparing current OAC use with non-use.

**Figure 1:**
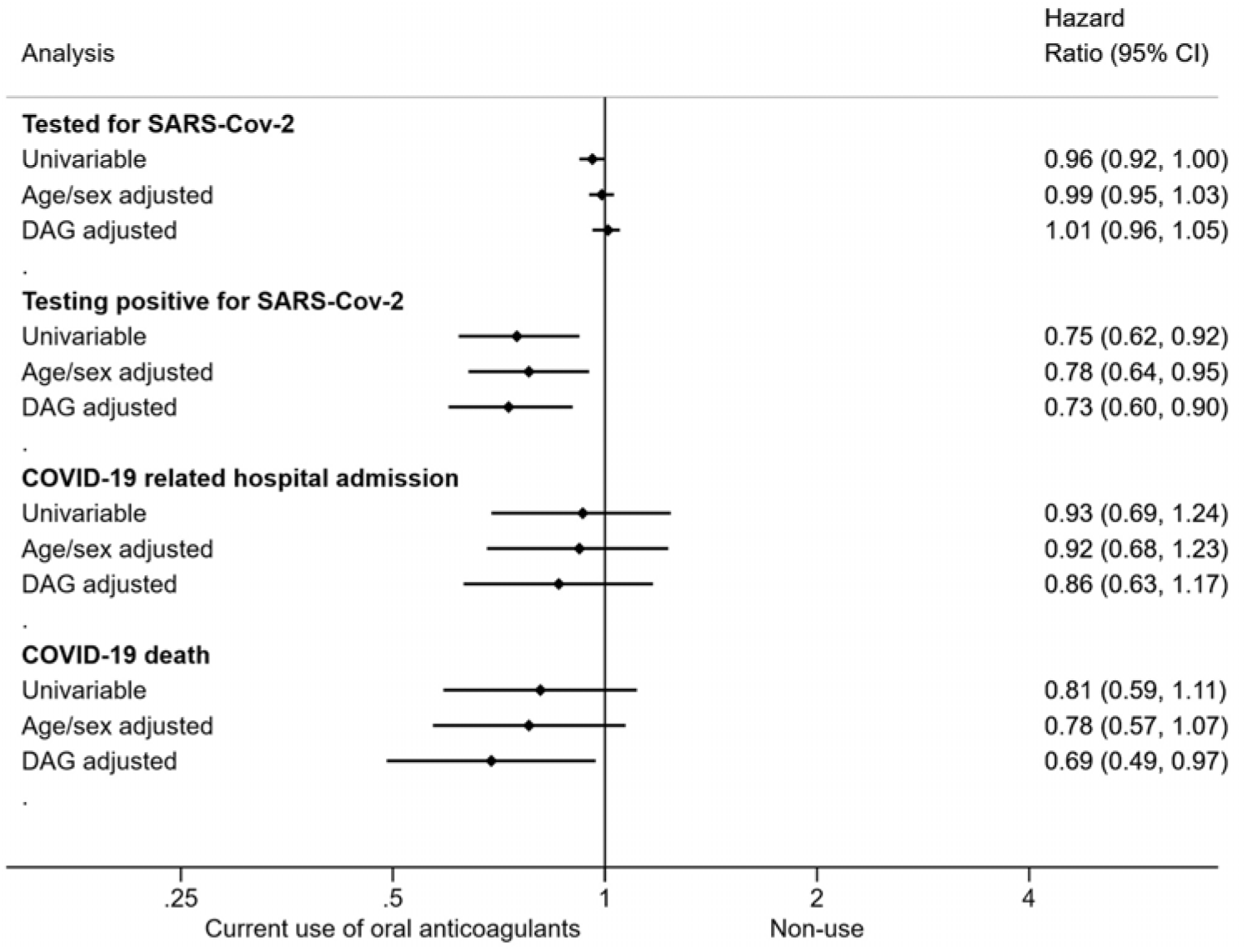
Hazard ratios of the association between current use of oral anticoagulants and COVID-19 related outcomes, versus non-use in people with atrial fibrillation who had a CHA□DS□-VASc score of 2.

#### Study 2: Risk of COVID-19 outcomes and non-COVID-19 death comparing warfarin with DOACs

We included 92,339 warfarin users and 280,407 DOAC users (Table 2). Median age was 79 years (IQR, 73 to 85) among warfarin users and 78 years (IQR, 71 to 84) among DOAC users. More men were warfarin users (60.6%) than DOAC users (56.2%).

Current warfarin users were more likely to be obese, former smokers and have a medical history of hypertension, heart failure, peripheral artery disease, venous thromboembolism, diabetes, chronic kidney disease than DOAC users. Current users were also less likely to have a prescription for oestrogen or oestrogen-like drugs, and antiplatelets, and to have had more primary care consultations and vaccinations than DOAC users.

Supplementary figure 6 presents time to each outcome in adjusted cumulative incidence plots. We observed a lower risk for all outcomes associated with current use of warfarin compared with current use of DOACs. For being tested for SARS-CoV-2, the unadjusted HR for current use of warfarin was 0.77 (95% CI, 0.76 to 0.79), with a DAG-adjusted HR of 0.80 (95% CI, 0.79 to 0.81) compared with current use of DOACs (Figure 2 & supplementary table 3). A lower risk of testing positive for SARS-CoV-2 [unadjusted HR: 0.69 (95% CI, 0.64 to 0.74); DAG adjusted HR: 0.73 (95% CI, 0.68 to 0.79)], COVID-19 related hospital admission [unadjusted HR: 0.77 (95% CI, 0.70 to 0.85); DAG adjusted HR: 0.75 (95% CI, 0.68 to 0.83)], and COVID-19 related deaths [unadjusted HR: 0.71 (95% CI, 0.64 to 0.79); DAG adjusted HR: 0.74 (95% CI, 0.66 to 0.83)] were observed comparing current use of warfarin with current use of DOACs. For non-COVID-19 deaths, the unadjusted HR was 0.81 (95% CI, 0.78 to 0.84); DAG adjusted HR: 0.79 (95% CI, 0.76 to 0.83) comparing current use of warfarin with current use of DOACs.

**Figure 2:**
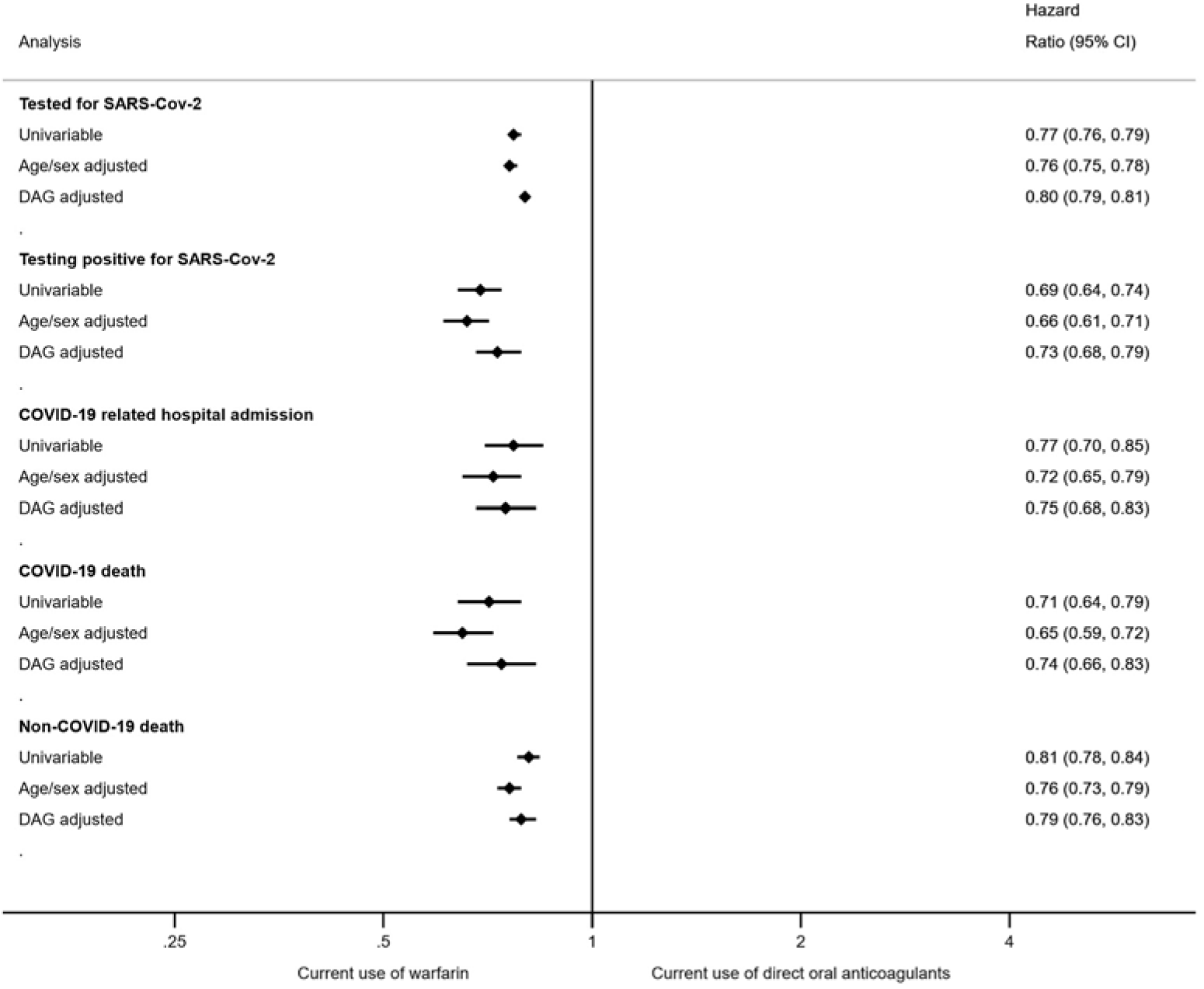
Hazard ratios of the association between current use of warfarin and COVID-19 related outcomes and non-COVID-19 deaths, versus direct oral anticoagulants in people with non-valvular atrial fibrillation.

In the *post-hoc* analyses investigating cause-specific deaths, we observed no difference in risk of deaths due to myocardial infarction, ischaemic stroke, venous thromboembolism, gastrointestinal bleeding and intracranial bleeding comparing current use of warfarin with current use of DOACs. However, the number of each specific outcome occurring was low, meaning power to observe any difference was limited. (Supplementary table 3).

### Sensitivity analyses

The results of all other sensitivity analyses were broadly similar to those of the main analyses (Supplementary tables 4-8, 10-12). Whilst there was no strong evidence of a different association between current use of OACs and testing positive for SARS-CoV-2 according to care home residence (Supplementary table 9), this comparison was underpowered with the DAG adjusted HR of 0.75 (95% CI, 0.60 to 0.93) in people not residing in care home and DAG adjusted HR of 1.01 (95% CI, 0.57 to 1.80) in people residing in care home (p-interaction: 0.33).

### Quantitative bias analysis

#### Study 1: Risk of COVID-19 outcomes comparing OAC use with non-use

We did not conduct bias analysis for being tested for SARS-CoV-2 comparing current use versus non-use of OAC as the observed hazard ratio was near null. To potentially move the observed point estimate (or upper bound of 95% CI) of the hazard ratio to a null bias-adjusted association an unmeasured confounder would need to be associated with either non-use, relative to OAC use or each outcome by at least RR 2.08 (1.46) for testing positive for SARS-CoV-2, 1.60 for COVID-19 related hospital admission, and 2.26 (1.21) for COVID-19 related mortality (E-value); and associated with both exposure and outcome (Cornfield condition) by at least 1.37 (1.11) for testing positive for SARS-CoV-2, 1.16 for COVID-19 hospital admission, and 1.45 (1.03) for COVID-19 related mortality (Supplementary table 13).

#### Study 2: Risk of COVID-19 outcomes comparing warfarin with DOACs

To potentially move the observed point estimate (upper bound of 95% CI) to a null bias-adjusted association, an unmeasured confounder would need to be associated with either receiving DOACs relative to warfarin, or each outcome (E-value) by at least 1.81 (1.77) for being tested for SARS-CoV-2, 2.08 (1.85) for testing positive for SARS-CoV-2, 2.00 (1.70) for COVID-19 related hospital admission, and 2.04 (1.70) for COVID-19 related death; and be associated with both exposure and outcome (Cornfield condition) by at least 1.25 (1.23) for being tested for SARS-CoV-2, 1.37 (1.27) for testing positive for SARS-CoV-2, 1.33 (1.20) for COVID-19 related hospital admission, 1.35 (1.20) for COVID-19 related death (Supplementary table 14).

An unmeasured confounder would need to be associated with both receiving DOACs, relative to warfarin, and each outcome (Cornfield condition), by associations ranging from 1.45 to 1.52 to move from the observed upper bound of the CI for the protective associations to bias-adjusted harmful association of HR 1.2.

## Discussion

### Principal findings

Based on routinely collected data, we observed a lower risk of testing positive for SARS-CoV-2 and COVID-19 related deaths associated with current OAC use, compared with non-use among people with AF who had a low baseline risk of stroke. We observed no difference in risk of being tested for SARS-CoV-2 between current OAC users and non-users, indicating that the lower risk of testing positive was unlikely due to the chance of being tested.

We also observed a lower risk of being tested for SARS-CoV-2, testing positive for SARS-CoV-2, COVID-19 related hospital admission, COVID-19 related deaths and non-COVID-19 death associated with current use of warfarin, compared with DOACs among people with non-valvular AF. These were consistently seen across all analyses.

Consideration needs to be given to whether these associations are causal, or due to other differences between comparison groups. There is no clear evidence of better or worse underlying health characteristics between current OAC users and non-users. The inverse associations we found in OAC users with AF and a low risk of stroke were specific to COVID-19 outcomes, with no protective association seen against having a COVID-19 test, which would support a possible causal association. OAC users had a reduced risk of receiving a positive test, as well as a reduced risk of severe COVID-19 outcomes, suggesting a lower risk of acquiring infection in this group. An experimental study suggested that direct factor Xa inhibitors may prevent SARS-CoV entry to human cells by preventing the spike protein cleavage into the S1 and S2 subunits^23^ but clinical evidence of the protective effects of OACs on SARS-CoV-2 infection is limited. Additionally, we considered that anticoagulation might inhibit the PCR for SARS-CoV-2. Some in-vivo studies showed that heparin inhibits different PCRs for other viruses,^24,25^ but the evidence of the inhibitory effect of OACs on PCRs for SARS-CoV-2 is lacking. Regardless, in many instances the outcome of a positive COVID-19 test reflects both infection and symptom severity leading to test seeking, the latter of which could have been influenced by anticoagulant activity. We also considered the non-causal explanation that health and risk behaviours may differ between OAC users and non-users and impact the probability of being infected with SARS-CoV-2 and severe COVID-19 outcomes. Although we cannot fully capture the behavioural differences between exposure groups in the database, we compared the smoking status, hazardous alcohol use and their receipts of flu vaccinations between exposure groups. It showed that OAC users (or warfarin users) are less likely to be current smokers, had less hazardous alcohol use and more likely to have had flu vaccination than non-users (or DOAC users) but the differences were small. Importantly, we adjusted for a range of confounders that made minimal impact to the estimates. No information was directly available on shielding behaviours during the pandemic, but we conducted a *post-hoc* subgroup analysis according to care home residence as people living in care homes were less likely to have behavioural differences to explain the findings. There was no difference between OAC users and non-users living in care homes, but this analysis was underpowered, limiting the interpretation of the findings. Further studies that can account for behavioural differences between groups are required to confirm the findings as we cannot rule this out as a possible contributor to our findings.

The protective associations seen for warfarin versus DOACs in all patients with AF are more surprising given the hypothesis of a possible harm with warfarin and generally more comorbidities among warfarin users than DOAC users. Also, our findings were non-specific, including an inverse association with receiving a COVID-19 test, and death from non-COVID causes. We tried to explore non-COVID deaths further to understand whether this finding was driven by possible therapeutic effects of warfarin, since others have also observed this association^26^ but there were too few outcomes to draw conclusions. Similar modestly greater health seeking and lower risk behaviour were seen amongst warfarin users compared with DOAC users, as noted in Study 1, though again, adjustment for these had little impact. Importantly, the results for warfarin do not suggest that it is associated with COVID-19 related harm.

We further performed quantitative bias analyses for both studies and found that an unmeasured confounder of moderate strength could potentially explain the observed associations. Replicating these findings using other large datasets and robust methods would be welcomed.

### Findings in Context

While the effects of COVID-19 may predispose patients to thromboembolic disease through severe illness, hypoxia or severe inflammatory response^27^, anticoagulation may have a role in preventing thrombotic events in patients with COVID-19. Recent studies investigating the potential effects of early initiation of prophylactic use of anticoagulation or therapeutic anticoagulation resulted in conflicting findings.^3–11,28–32^ In line with our research, most studies reported beneficial effects of anticoagulation on severe COVID-19 outcomes including mortality or ICU admission.

Seven studies^4,5,9,10,12–14^ focusing on prehospital use of anticoagulants and one study^29^ focusing on therapeutic use of anticoagulants found no difference in risk of mortality, mechanical ventilation or acute distress respiratory syndrome. Notably, some were of small sample size^4,12–14^, with unclear exposure definition^5^, classifying patients who initiated therapeutic anticoagulation on day 3 or later after ICU admission in the control group^29^ or used composite outcomes^9,10^, limiting the interpretation of the results. Two cohort studies showed a higher risk of admission to ICU, intubation, or death associated with anticoagulants in patients with COVID-19 disease, compared with non-use, but the results were possibly due to confounding by indication.^8,31^

### Strengths and limitations

The greatest strength of this study was the power enabling us to examine the association between OACs and various COVID-19 related outcomes as our dataset included medical records from 24 million individuals. To our knowledge, this is also the first population-based study comparing risk of COVID-related outcomes and cause-specific deaths between warfarin and DOACs. We also conducted quantitative bias analyses to explore the impact of unmeasured confounding to our observed results, supporting our interpretation. The breadth of data available in primary care allows us to account for a wide range of potential confounders. We pre-specified our analysis plan and have shared all analytical code.

We recognise possible limitations. First, we could not eliminate residual confounding. Whilst differences in health behaviours and shielding between groups may partly explain our results, more studies are required to confirm these findings. Second, we do not know whether patients took the medications as prescribed. We are also not able to capture any anticoagulant use during hospitalisation. Low molecular weight heparin or unfractionated heparin might be given during hospitalisation for OAC untreated patients with severe COVID-19 disease to prevent venous thromboembolism, merely leading to an underestimation of the effect without accounting for the anticoagulation use during hospitalisation. Third, there may be misclassification in ascertaining AF using diagnostic codes alone when deriving the study population. Some recorded AF might resolve at the study start and thus not require anticoagulants. However, this is considered to be less common^33^ and is unlikely to substantially bias our results. Although we attempted to reduce confounding by limiting the study cohort to people who had a threshold CHA□DS□-VASc score to be prescribed anticoagulants in Study 1, results may not be generalisable to all patients with AF. In particular, women and people with stroke, transient ischaemic attack or venous thromboembolism may be under-represented in this study given that these alone would have led to a CHA□DS□-VASc score of 2, so any additional risk factors would mean therefore would be excluded from the exposed group.

## Policy Implications

Further studies are needed to confirm our findings of an inverse association between OACs and severe COVID-19 related outcomes in people with AF at the threshold for OAC treatment, and to establish causality; if confirmed to be a causal effect, this could be of significant clinical importance, particularly as the older age and comorbidities in this group are independent risk factors of severe COVID-19 outcomes.

This study also shows no evidence of a higher risk of COVID-19 related outcomes associated with warfarin versus DOACs, providing reassurance about the safety of warfarin use during the COVID-19 pandemic. Choice of routine anticoagulant therapy represents a complex balance of expected risks, benefits and patient preference; we do not recommend changes to ongoing anticoagulant therapy based on these results.

## Conclusions

We showed a lower risk of testing positive for SARS-CoV-2 and COVID-19 related deaths associated with routinely prescribed OACs in people with AF who had a threshold of being treated with OAC based on risk of stroke. This might be explained by a potential effect of OACs on preventing severe COVID-19 outcomes before hospitalisation, or more cautious behaviours and environmental factors reducing the risk of SARS-CoV-2 infection in those taking OACs.

There is no evidence of a higher risk of severe COVID-19 outcomes associated with warfarin compared with DOACs, providing reassurance about the safety of warfarin use among patients with indications for anticoagulation in the context of the COVID-19 pandemic.

## Data Availability

All codelists for identifying exposures, covariates and outcomes are openly shared at https://codelists.opensafely.org/ for inspection and re-use. All study analyses were pre-planned unless otherwise stated. All code for data management and analyses in addition to the pre-specified protocol (https://github.com/opensafely/anticoagulants-research/blob/master/protocol/Protocol_%20Anticoag%20OpenSAFELY_v3.docx) are archived at: https://github.com/opensafely/anticoagulants-research.

## Acknowledgements

We are very grateful for all the support received from the TPP Technical Operations team throughout this work; for generous assistance from the information governance and database teams at NHS England / NHSX.

## Copyright/licence for publication

The Corresponding Author has the right to grant on behalf of all authors and does grant on behalf of all authors, a worldwide licence to the Publishers and its licensees in perpetuity, in all forms, formats and media (whether known now or created in the future), to i) publish, reproduce, distribute, display and store the Contribution, ii) translate the Contribution into other languages, create adaptations, reprints, include within collections and create summaries, extracts and/or, abstracts of the Contribution, iii) create any other derivative work(s) based on the Contribution, iv) to exploit all subsidiary rights in the Contribution, v) the inclusion of electronic links from the Contribution to third party material where-ever it may be located; and, vi) licence any third party to do any or all of the above.

## Transparency declaration

The lead author^*^ affirms that this manuscript is an honest, accurate, and transparent account of the study being reported; that no important aspects of the study have been omitted; and that any discrepancies from the study as planned (and, if relevant, registered) have been explained. *The manuscript’s guarantor.

## Information Governance

NHS England is the data controller; TPP is the data processor; and the key researchers on OpenSAFELY are acting on behalf of NHS England. This implementation of OpenSAFELY is hosted within the TPP environment which is accredited to the ISO 27001 information security standard and is NHS IG Toolkit compliant;^34,35^ patient data has been pseudonymised for analysis and linkage using industry standard cryptographic hashing techniques; all pseudonymised datasets transmitted for linkage onto OpenSAFELY are encrypted; access to the platform is via a virtual private network (VPN) connection, restricted to a small group of researchers, their specific machine and IP address; the researchers hold contracts with NHS England and only access the platform to initiate database queries and statistical models; all database activity is logged; only aggregate statistical outputs leave the platform environment following best practice for anonymisation of results such as statistical disclosure control for low cell counts.^36^ The OpenSAFELY research platform adheres to the data protection principles of the UK Data Protection Act 2018 and the EU General Data Protection Regulation (GDPR) 2016. In March 2020, the Secretary of State for Health and Social Care used powers under the UK Health Service (Control of Patient Information) Regulations 2002 (COPI) to require organisations to process confidential patient information for the purposes of protecting public health, providing healthcare services to the public and monitoring and managing the COVID-19 outbreak and incidents of exposure.^37^ Taken together, these provide the legal bases to link patient datasets on the OpenSAFELY platform. GP practices, from which the primary care data are obtained, are required to share relevant health information to support the public health response to the pandemic, and have been informed of the OpenSAFELY analytics platform.

## Guarantor

AYSW/BG/ID are guarantors.

## Contributorship

Contributions are as follows:

Conceptualization LS BG ID;

Data curation CB JP JC SH SB DE PI CM;

Formal Analysis AYSW JB;

Funding acquisition BG LS;

Information governance AM BG CB;

Methodology ID AYSW LT AS EP WE KW KB CTR EW SJWE LS JB CM AJW BM SB RE BG;

Disease category conceptualisation and codelists AYSW LT WE BM CM AJW RC AS CTR PI SB DE CB JC HC KB AM ID HM RM HF RE;

Ethics approval HC EW LS BG;

Project administration AYSW BM CM AS AJW CTR WH CB SB AM LS BG;

Resources BG LS;

Software SB DE PI AJW CM CB JC;

Supervision ID BG;

Visualisation AYSW JB;

Writing (original draft) AYSW ID JB;

All authors were involved in design and conceptual development and reviewed and approved the final manuscript.

## Licence statement

I, the Submitting Author has the right to grant and does grant on behalf of all authors of the Work (as defined in the below author licence), an exclusive licence and/or a non-exclusive licence for contributions from authors who are: i) UK Crown employees; ii) where BMJ has agreed a CC-BY licence shall apply, and/or iii) in accordance with the terms applicable for US Federal Government officers or employees acting as part of their official duties; on a worldwide, perpetual, irrevocable, royalty-free basis to BMJ Publishing Group Ltd (“BMJ”) its licensees and where the relevant Journal is co-owned by BMJ to the co-owners of the Journal, to publish the Work in Annals of the Rheumatic Diseases and any other BMJ products and to exploit all rights, as set out in our licence.

The Submitting Author accepts and understands that any supply made under these terms is made by BMJ to the Submitting Author unless you are acting as an employee on behalf of your employer or a postgraduate student of an affiliated institution which is paying any applicable article publishing charge (“APC”) for Open Access articles. Where the Submitting Author wishes to make the Work available on an Open Access basis (and intends to pay the relevant APC), the terms of reuse of such Open Access shall be governed by a Creative Commons licence – details of these licences and which Creative Commons licence will apply to this Work are set out in our licence referred to above.

## Abbreviation

AF: atrial fibrillation
CI: confidence interval
COVID-19: coronavirus disease 2019
DAG: Directed Acyclic Graph
DOAC: direct oral anticoagulants
HR: Hazard ratio
OAC: oral anticoagulants
NHS: National Health Service

