## Supplementary for "Association between oral anticoagulants and COVID-19 related outcomes: two cohort studies"

Deviations from pre-specified protocol, with reasons

We noted that people who were prescribed and not prescribed an OAC could have important clinical differences which might bias comparisons between them. To explore this as a pre-specified analysis, we further matched each identified current OAC user to up to 10 people from the general population (regardless of an AF diagnosis) based on age, sex, and general practice on study start date. The same exclusion criteria applied to the AF population were also applied. In addition, people who had no GP visit within one year before study start date or had any OAC prescription within 4 months before study start date were excluded. However, after matching, we observed very large differences in baseline characteristics between the OAC exposed group and the comparison group from general population (see Supplementary table 1), particularly on the proportions of ischaemic stroke and venous thromboembolism which are components of CHA₂DS₂-VASc score. This is because people with a CHA₂DS₂-VASc of 2 would tend to have a low prevalence of VTE and stroke, since these comorbidities would generally result in a CHA₂DS₂-VASc higher than 2, when age and other factors were added in. Given these large, conditional differences between groups, we determined any comparisons between them would be subject to substantial confounding bias and would likely be uninterpretable. This analysis was therefore not considered further in this study.

### **Supplementary table 1. Demographic and Clinical Characteristics comparing current use in people with atrial fibrillation with general population**

|  | **Matched general population** | **Current use** |
| --- | --- | --- |
| **Total** | 511,361 | 51,784 |
| **Age as of 1^st^ Mar2020** |  |  |
| 18-<40 | 1,247 (0.2) | 126 (0.2) |
| 40-<50 | 7,491 (1.5) | 753 (1.5) |
| 50-<60 | 46,100 (9.0) | 4,642 (9.0) |
| 60-<70 | 155,854 (30.5) | 15,673 (30.3) |
| 70-<80 | 223,949 (43.8) | 22,487 (43.4) |
| 80+ | 76,720 (15.0) | 8,103 (15.6) |
| Median, IQR | 71 (66-75) | 71 (66-75) |
| **Sex** |  |  |
| Female | 107,444 (21.0) | 10,819 (20.9) |
| **Body mass index** |  |  |
| <18.5 | 5,118 (1.0) | 496 (1.0) |
| 18.5-24.9 | 133,032 (26.0) | 11,667 (22.5) |
| 25-29.9 | 198,691 (38.9) | 17,751 (34.3) |
| 30-34.9 | 95,352 (18.6) | 10,984 (21.2) |
| 35-39.9 | 29,518 (5.8) | 4,720 (9.1) |
| 40+ | 11,022 (2.2) | 2,851 (5.5) |
| Missing | 38,628 (7.6) | 3,315 (6.4) |
| **Ethnicity** |  |  |
| White | 375,297 (73.4) | 37,729 (72.9) |
| Mixed | 1,635 (0.3) | 102 (0.2) |
| Asian/Asian British | 10,975 (2.1) | 506 (1.0) |
| Black | 4,039 (0.8) | 182 (0.4) |
| Other | 3,080 (0.6) | 217 (0.4) |
| Missing | 116,335 (22.8) | 13,048 (25.2) |
| **Index of Multiple Deprivation** |  |  |
| 1 (least deprived) | 102,420 (20.0) | 11,242 (21.7) |
| 2 | 103,620 (20.3) | 10,914 (21.1) |
| 3 | 101,931 (19.9) | 10,371 (20.0) |
| 4 | 101,868 (19.9) | 9,818 (19.0) |
| 5 (most deprived) | 101,522 (19.9) | 9,439 (18.2) |
| **Smoking status** |  |  |
| Never | 182,618 (35.7) | 18,776 (36.3) |
| Former | 273,612 (53.5) | 29,072 (56.1) |
| Current | 53,977 (10.6) | 3,847 (7.4) |
| Missing | 1,154 (0.2) | 89 (0.2) |
| **Hazardous alcohol use** | 55,771 (10.9) | 6,435 (12.4) |
| **Comorbidities** |  |  |
| Hypertension | 249,883 (48.9) | 21,613 (41.7) |
| Heart Failure | 15,817 (3.1) | 5,570 (10.8) |
| Myocardial infarction | 32,387 (6.3) | 1,597 (3.1) |
| Peripheral arterial disease | 14,462 (2.8) | 419 (0.8) |
| Stroke/transient ischaemic attack | 36,184 (7.1) | 1,023 (2.0) |
| Venous thromboembolism | 12,948 (2.5) | 544 (1.1) |
| Diabetes |  |  |
| Controlled  (HbA1c < 58 mmols/mol) | 76,745 (15.0) | 4,209 (8.1) |
| Uncontrolled  (HbA1c ≥ 58 mmols/mol) | 32,389 (6.3) | 2,073 (4.0) |
| HbA1c not measured | 1,309 (0.3) | 70 (0.1) |
| COPD | 44,475 (8.7) | 5,151 (9.9) |
| Other respiratory diseases | 16,602 (3.2) | 2,081 (4.0) |
| Cancer | 70,747 (13.8) | 7,832 (15.1) |
| Immunosuppression | 3,507 (0.7) | 434 (0.8) |
| Chronic kidney disease | 65,497 (12.8) | 8,291 (16.0) |
| **Primary care consultations** |  |  |
| Median, IQR | 6 (3-11) | 10 (5-16) |
| Min, Max | 1, 386 | 0, 198 |
| **A&E attendance** |  |  |
| Median, IQR | 0 (0-0) | 0 (0-1) |
| Min, Max | 0, 70 | 0, 54 |
| **Vaccination** |  |  |
| Flu | 335,861 (65.7) | 39,115 (75.5) |
| **Medications** |  |  |
| Oestrogen/ oestrogen-like drugs | 4,207 (0.8) | 352 (0.7) |
| Antiplatelets | 105,386 (20.6) | 2,264 (4.4) |

### **Supplementary figure 1.** Follow-up diagrams

(a) Study 1: Risk of COVID-19 outcomes comparing oral anticoagulant use and non-use

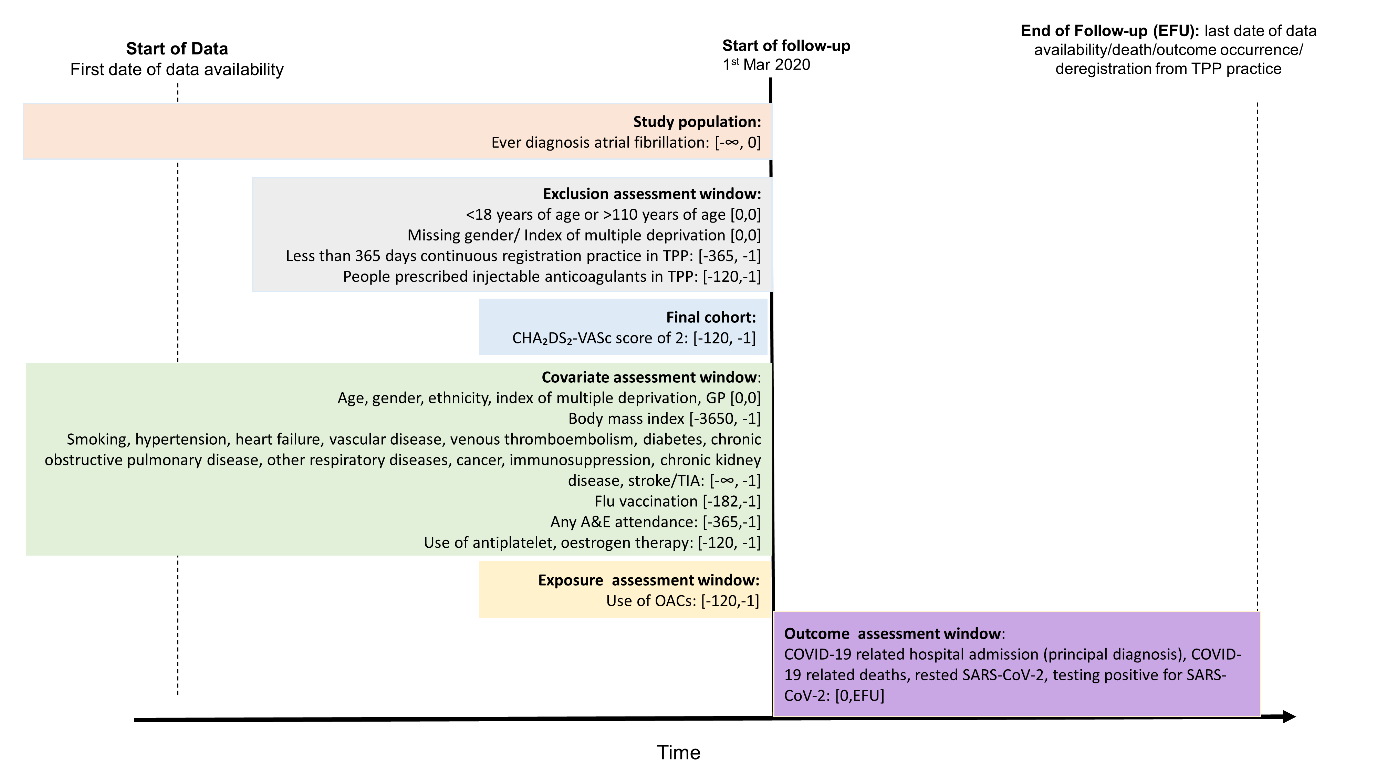

(b) Study 2: Risk of COVID-19 outcomes and non-COVID-death comparing warfarin and direct oral anticoagulants

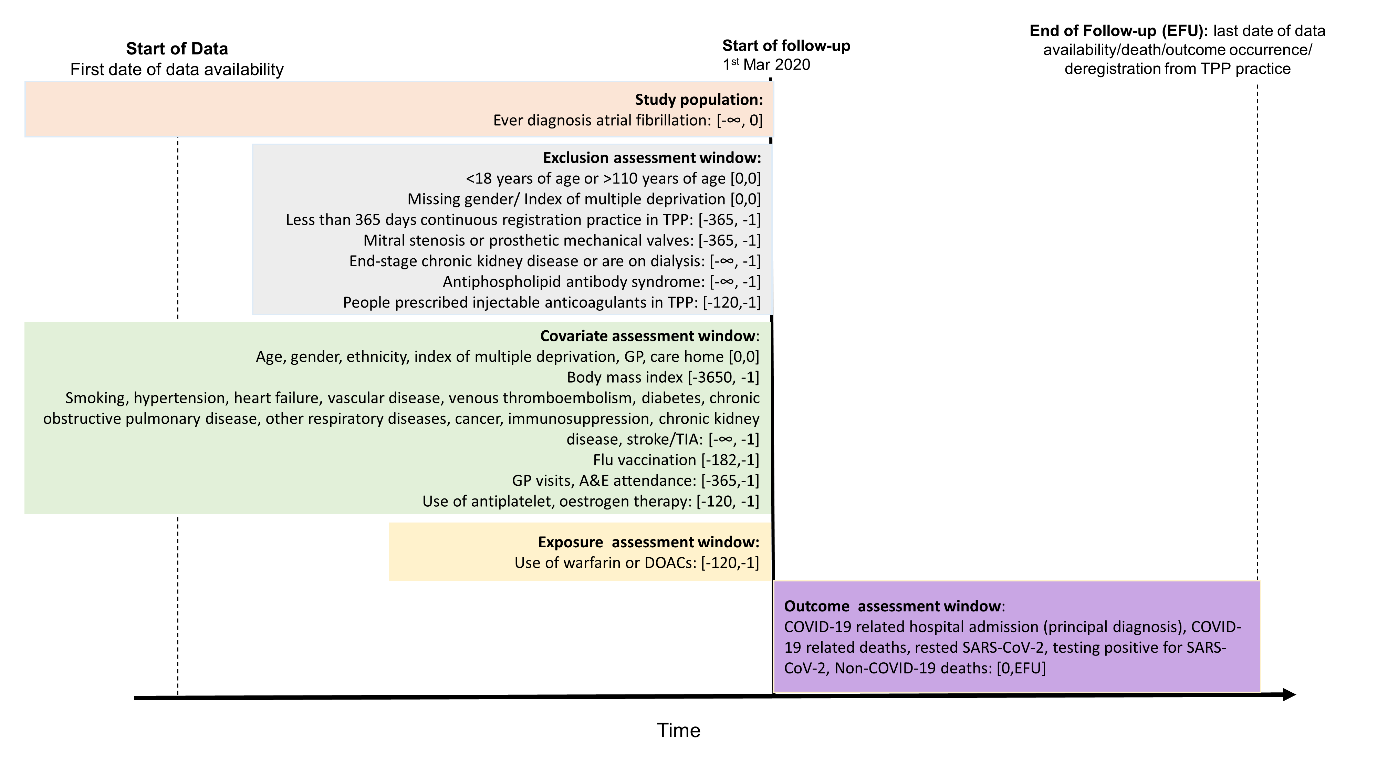

### **Supplementary figure 2.** Directed Acyclic Graph for Study 1 investigating COVID-19 related outcomes comparing current use of oral anticoagulants and non-use

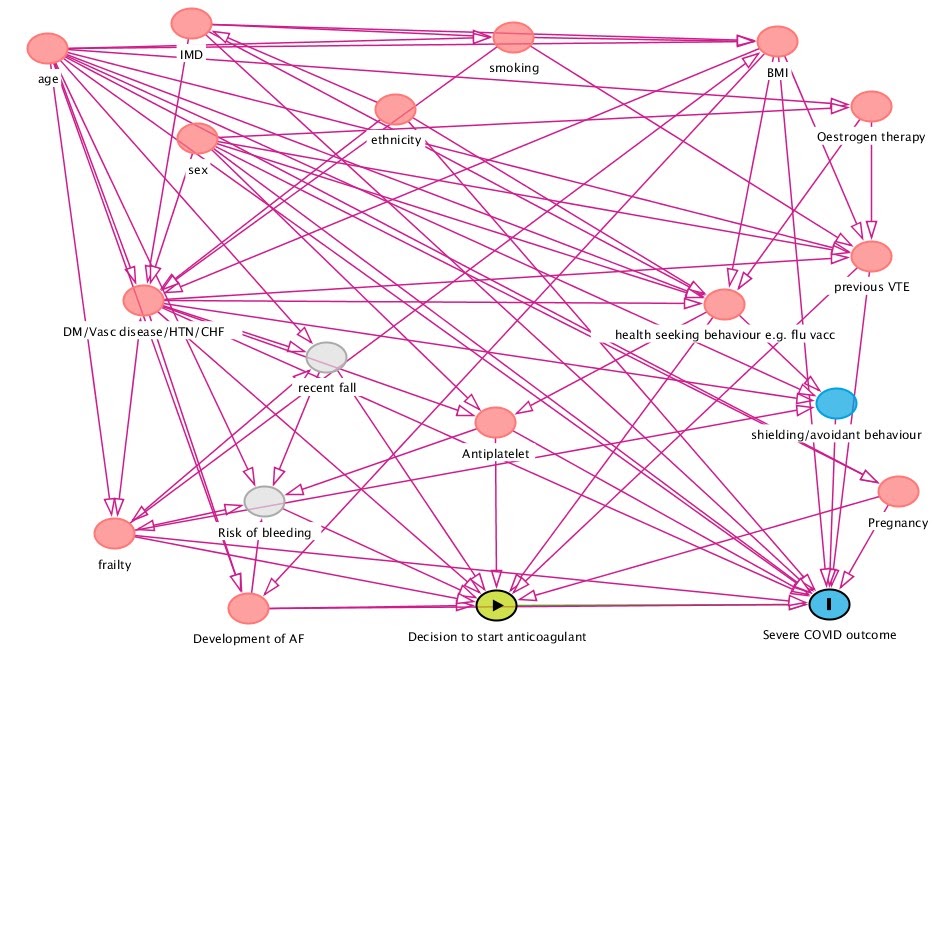

### **Supplementary figure 3.** Directed Acyclic Graph for Study 2 investigating COVID-19 related outcomes and non-COVID-19 deaths comparing current use of warfarin and direct oral anticoagulants

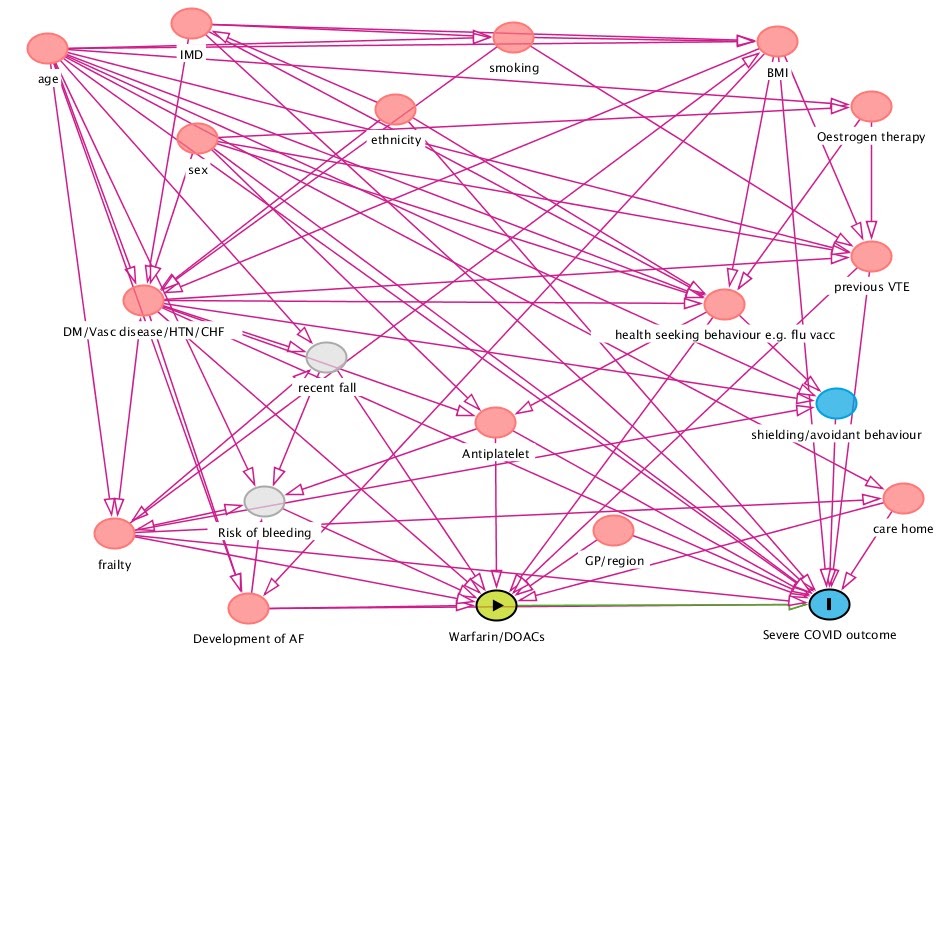

Supplementary figure 4. flowchart of inclusion of participants

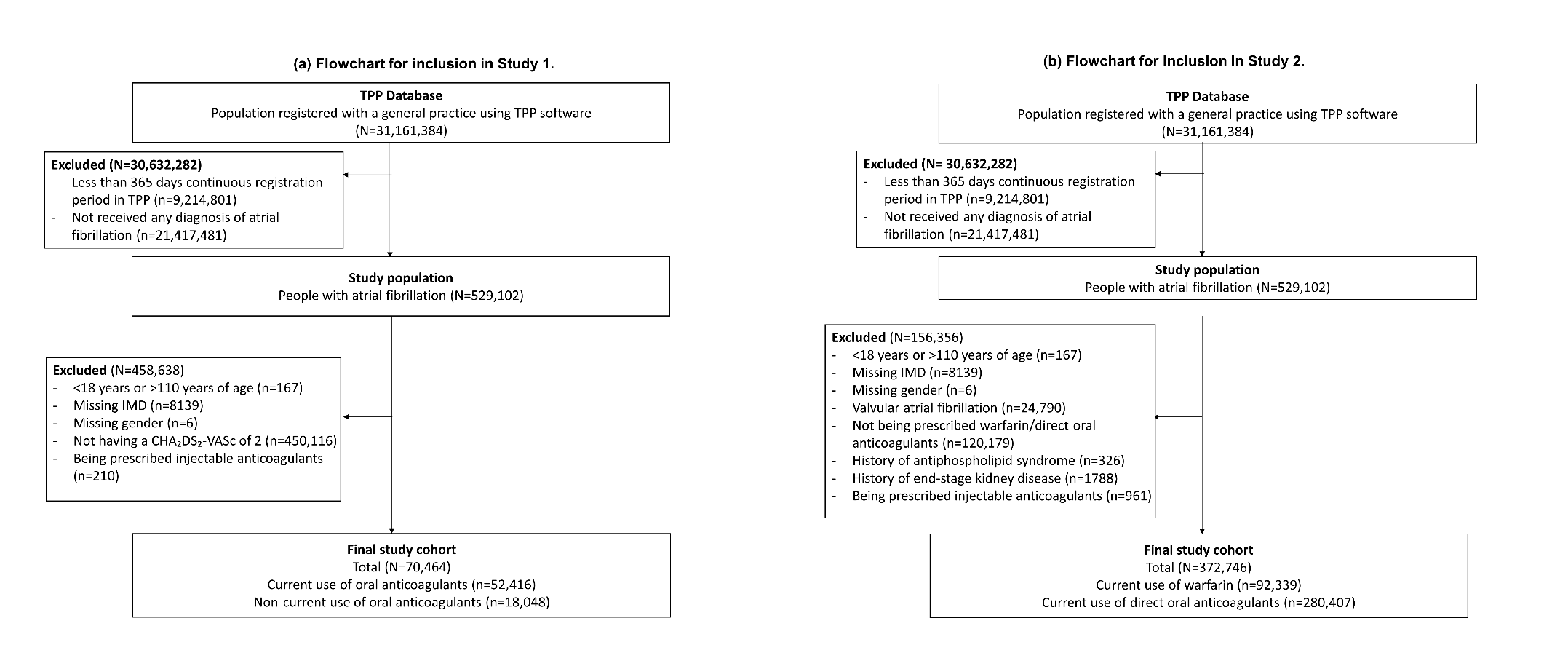

### Supplementary figure 5. Time to COVID-19 outcomes in adjusted cumulative incidence plots in Study 1

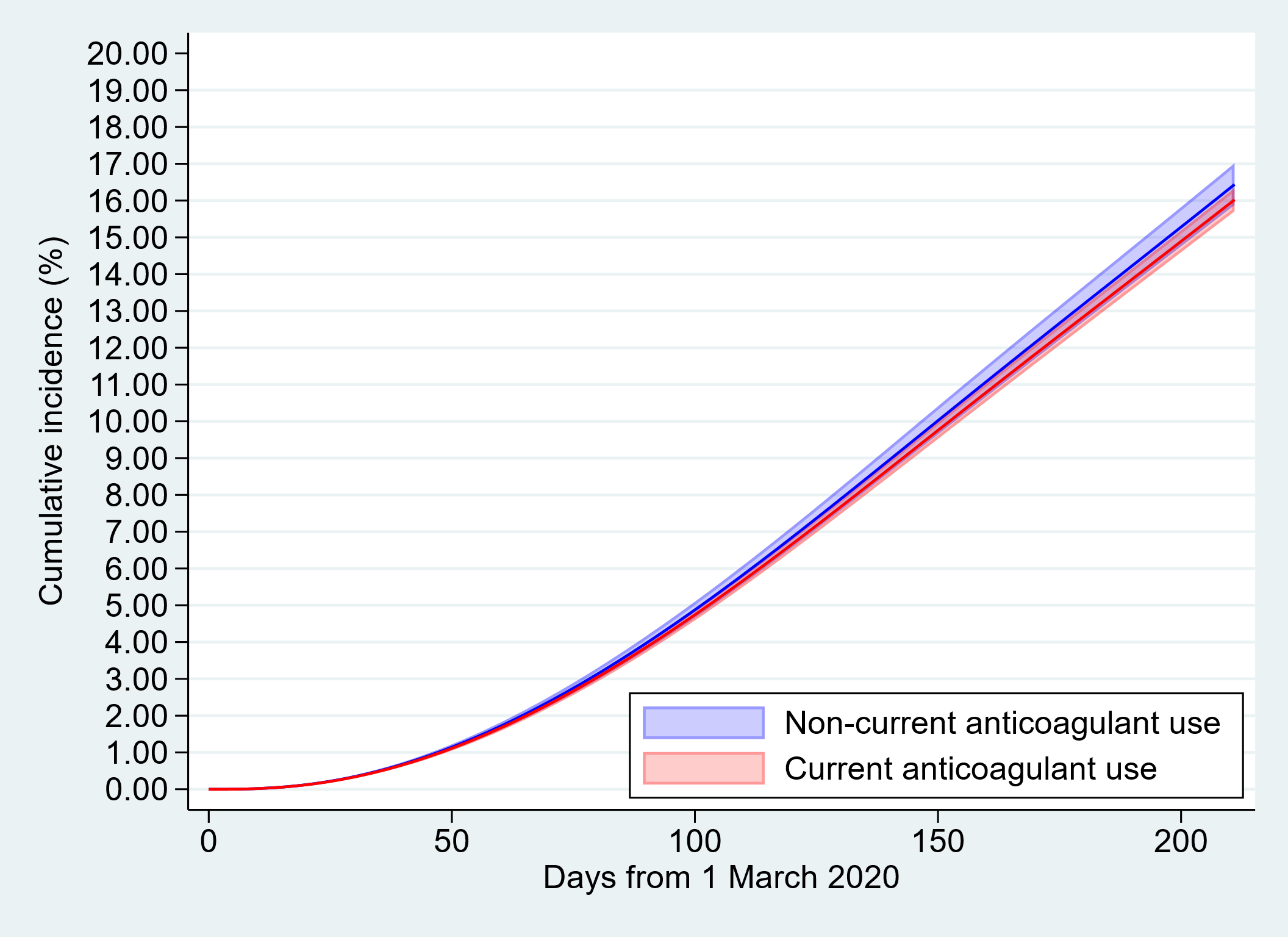
 **(a) Time to being tested for SARS-CoV-2**

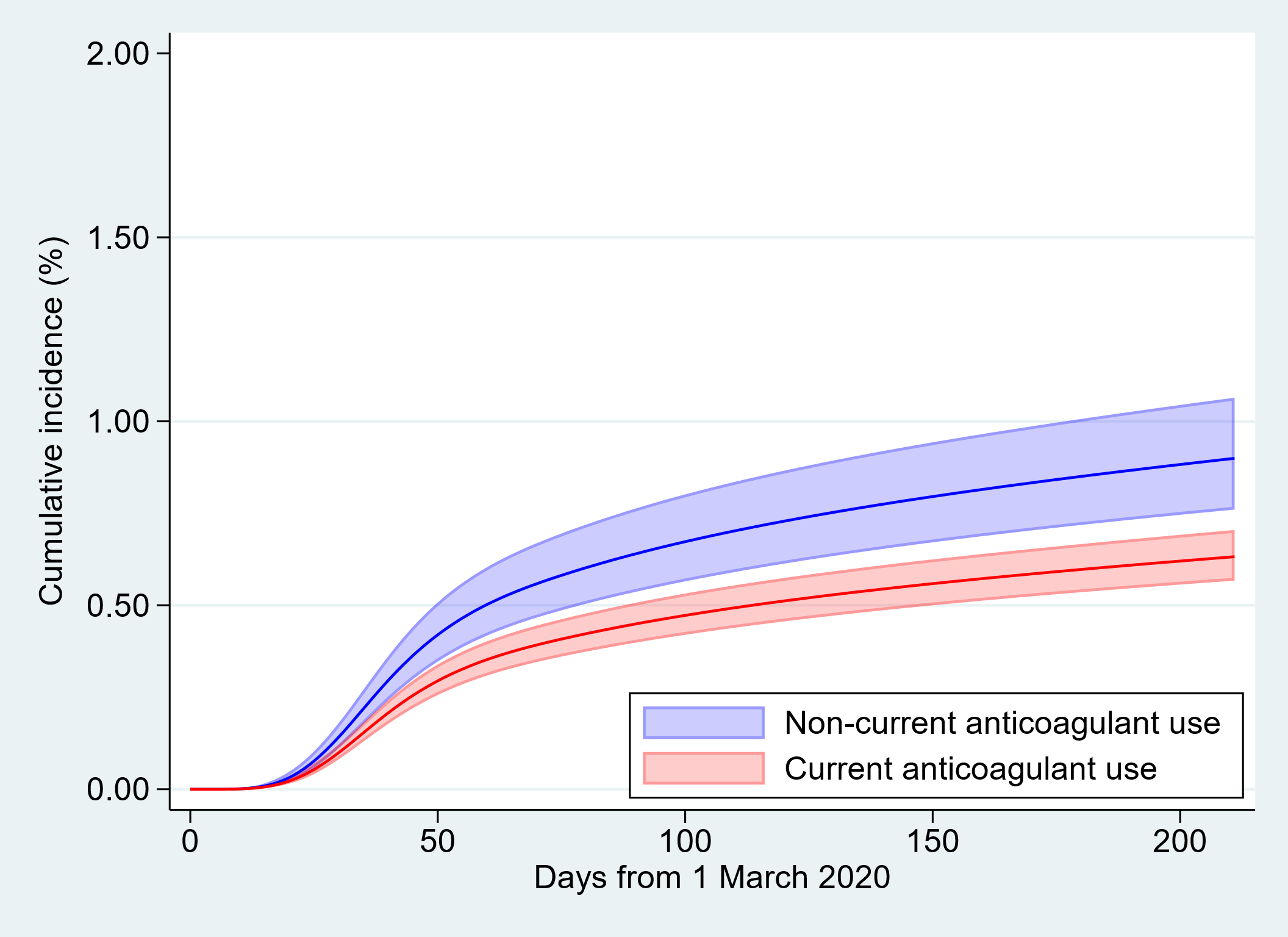
 **(b) Time to testing positive for SARS-CoV-2**

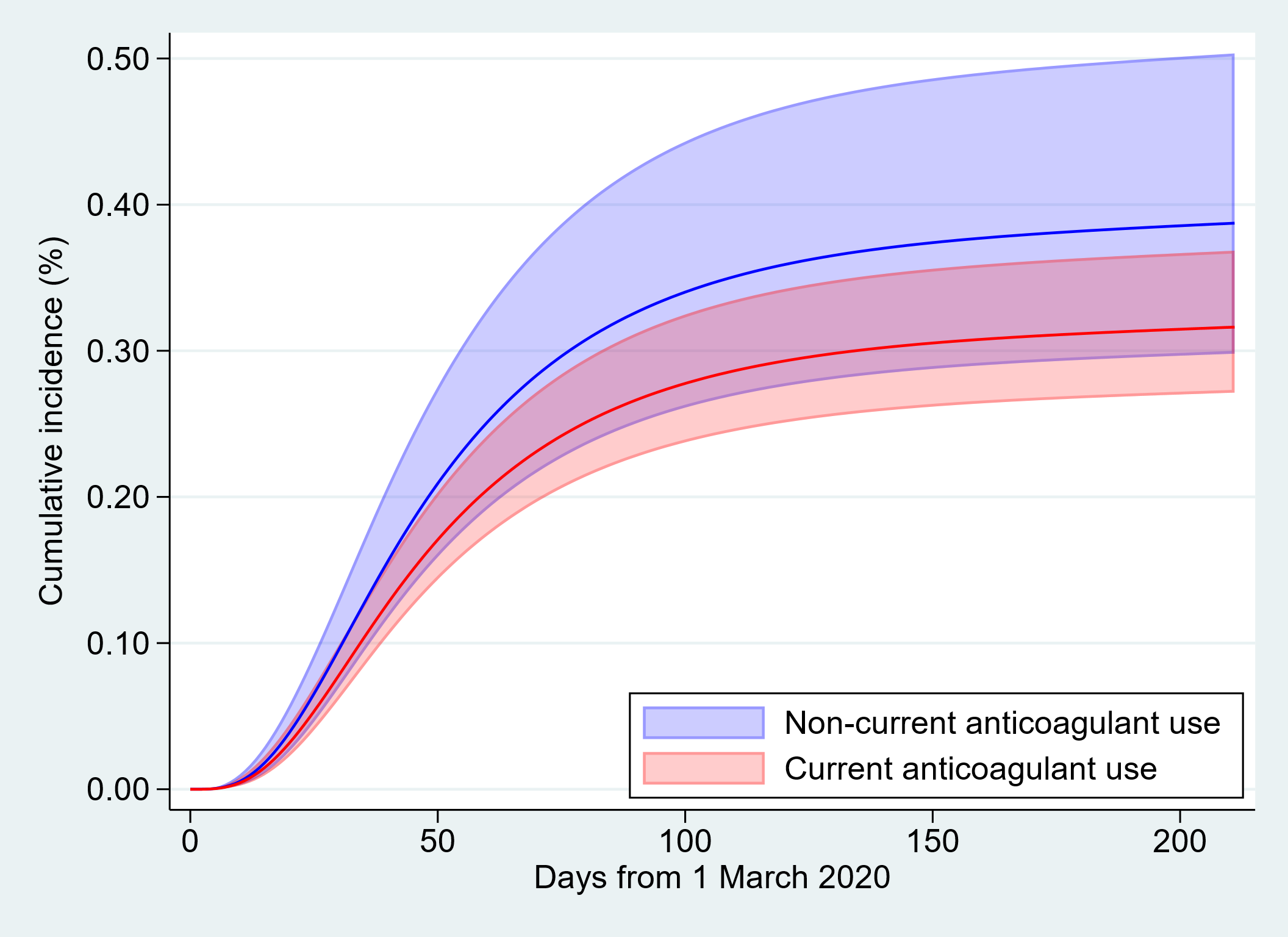
 **(c) Time to COVID-19 related hospital admission**

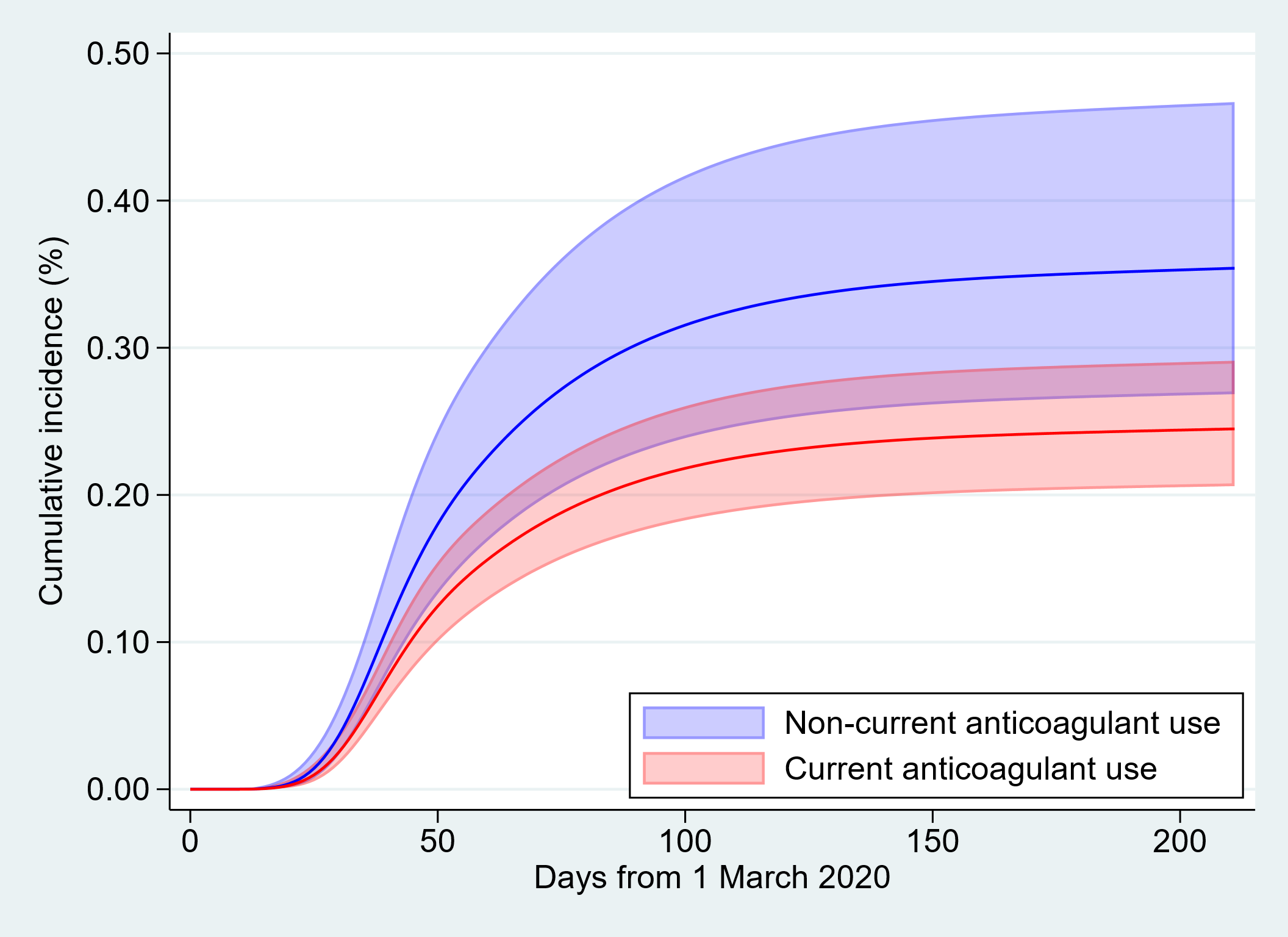
 **(d) Time to COVID-19 related deaths**

These figures present cumulative incidence and mortality predicted from a Royston-Parmar model including all covariates from the fully-adjusted Cox model, with the baseline hazard parametrized as a 2-degrees-of-freedom cubic spline for COVID-19 related hospital admissions, 3-degrees-of-freedom cubic spline for being tested for SARS-CoV-2, and testing positive for SARS-CoV-2, and COVID-19 related deaths; predictions standardized to the covariate distribution of the exposure group.

### **Supplementary table 2.** Results in the DAG and fully adjusted models comparing current use of oral anticoagulants with non-use in Study 1.

|  | **Number of events** | **Total person-weeks** | **Rate per 1,000** | **Unadjusted** | | **Age/Sex Adjusted** | | **DAG Adjusted*** | | **Fully adjusted^a^** | | |
| --- | --- | --- | --- | --- | --- | --- | --- | --- | --- | --- | --- | --- |
|  |  |  |  | HR | 95% CI | HR | 95% CI | HR | 95% CI | | HR | 95% CI |
| **Tested for SARS-CoV-2** |  |  |  |  |  |  |  |  |  | |  |  |
| non-use | 2922 | 500852 | 5.83 | 1.00 (ref) |  | 1.00 (ref) |  | 1.00 (ref) |  | | 1.00 (ref) |  |
| current use | 8248 | 1474160 | 5.60 | 0.96 | 0.92 - 1.00 | 0.99 | 0.95 - 1.03 | 1.01 | 0.96 - 1.05 | | 0.97 | 0.92 - 1.02 |
| **Testing positive for SARS-CoV-2** | |  |  |  |  |  |  |  |  | |  |  |
| non-use | 150 | 531336 | 0.28 | 1.00 (ref) |  | 1.00 (ref) |  | 1.00 (ref) |  | | 1.00 (ref) |  |
| current use | 331 | 1556683 | 0.21 | 0.75 | 0.62 - 0.92 | 0.78 | 0.64 - 0.95 | 0.73 | 0.60 - 0.90 | | 0.68 | 0.55 - 0.85 |
| **COVID-19 related hospital admission^b^** | |  |  |  |  |  |  |  |  | |  |  |
| non-use | 62 | 532501 | 0.12 | 1.00 (ref) |  | 1.00 (ref) |  | 1.00 (ref) |  | | 1.00 (ref) |  |
| current use | 168 | 1558848 | 0.11 | 0.93 | 0.69 - 1.24 | 0.92 | 0.68 - 1.23 | 0.86 | 0.63 - 1.17 | | 0.76 | 0.55 - 1.06 |
| **COVID-19 death^b,c^** |  |  |  |  |  |  |  |  |  | |  |  |
| non-use | 55 | 533397 | 0.10 | 1.00 (ref) |  | 1.00 (ref) |  | 1.00 (ref) |  | | 1.00 (ref) |  |
| current use | 130 | 1560931 | 0.08 | 0.81 | 0.59 - 1.11 | 0.78 | 0.57 - 1.07 | 0.69 | 0.49 - 0.97 | | 0.68 | 0.47 - 0.98 |

*Adjusted for age, sex, obesity, smoking, hypertension, heart failure, myocardial infarction, peripheral arterial disease, stroke/transient ischemic attack, venous thromboembolism, diabetes, flu vaccination, antiplatelet use, oestrogen and oestrogen-like therapy use, and Index of Multiple Deprivation.

^a^Adjusted for age, sex, obesity, smoking, hypertension, heart failure, myocardial infarction, peripheral arterial disease, stroke/transient ischemic attack, venous thromboembolism, diabetes, flu vaccination, antiplatelet use, oestrogen and oestrogen-like therapy use, Index of Multiple Deprivation, chronic obstructive pulmonary disease, other respiratory diseases, cancer, immunosuppression, chronic kidney disease, general practice attendance and A&E attendance and stratified on general practice.

^b^For outcomes of COVID-19 related hospital admission and COVID-19 death, we classified people with a diabetes diagnosis but not having HbA1c measures in the past year as uncontrolled diabetes in DAG adjusted and fully adjusted models.as the parameter for not having HbA1c measures did not converge and people with a diabetes diagnosis but not having HbA1c measures in the past year, are likely to have uncontrolled diabetes due to their potential lack of monitoring and management of diabetes.

^c^Due to low event count for parameters of stroke/transient ischaemic attack and oestrogen use, they did not converge in the model and were dropped from the main analysis.

Supplementary figure 6. Time to COVID-19 outcomes and non-COVID-19 death in adjusted cumulative incidence plots in Study 2
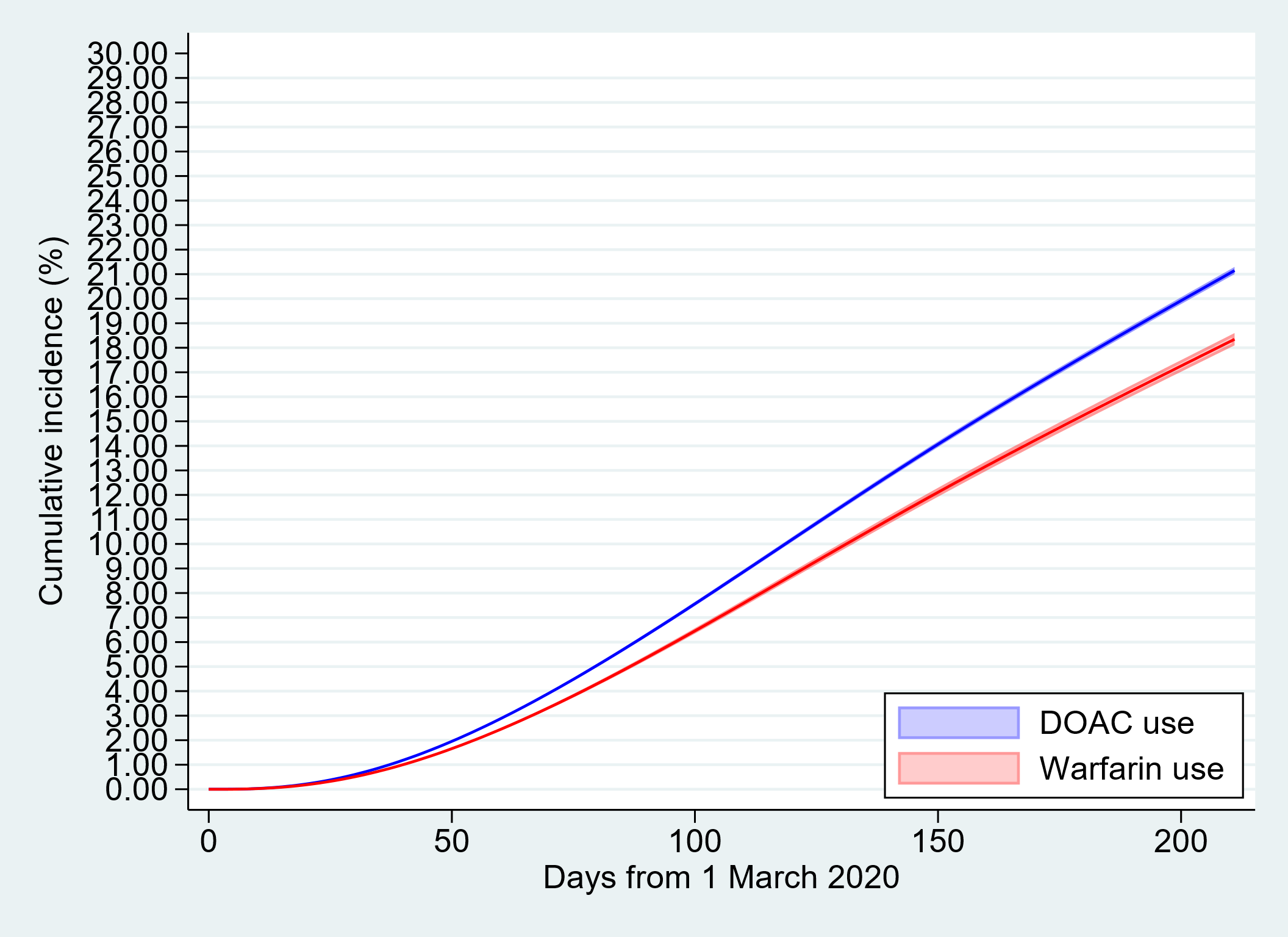
 **(a) Time to being tested for SARS-CoV-2**
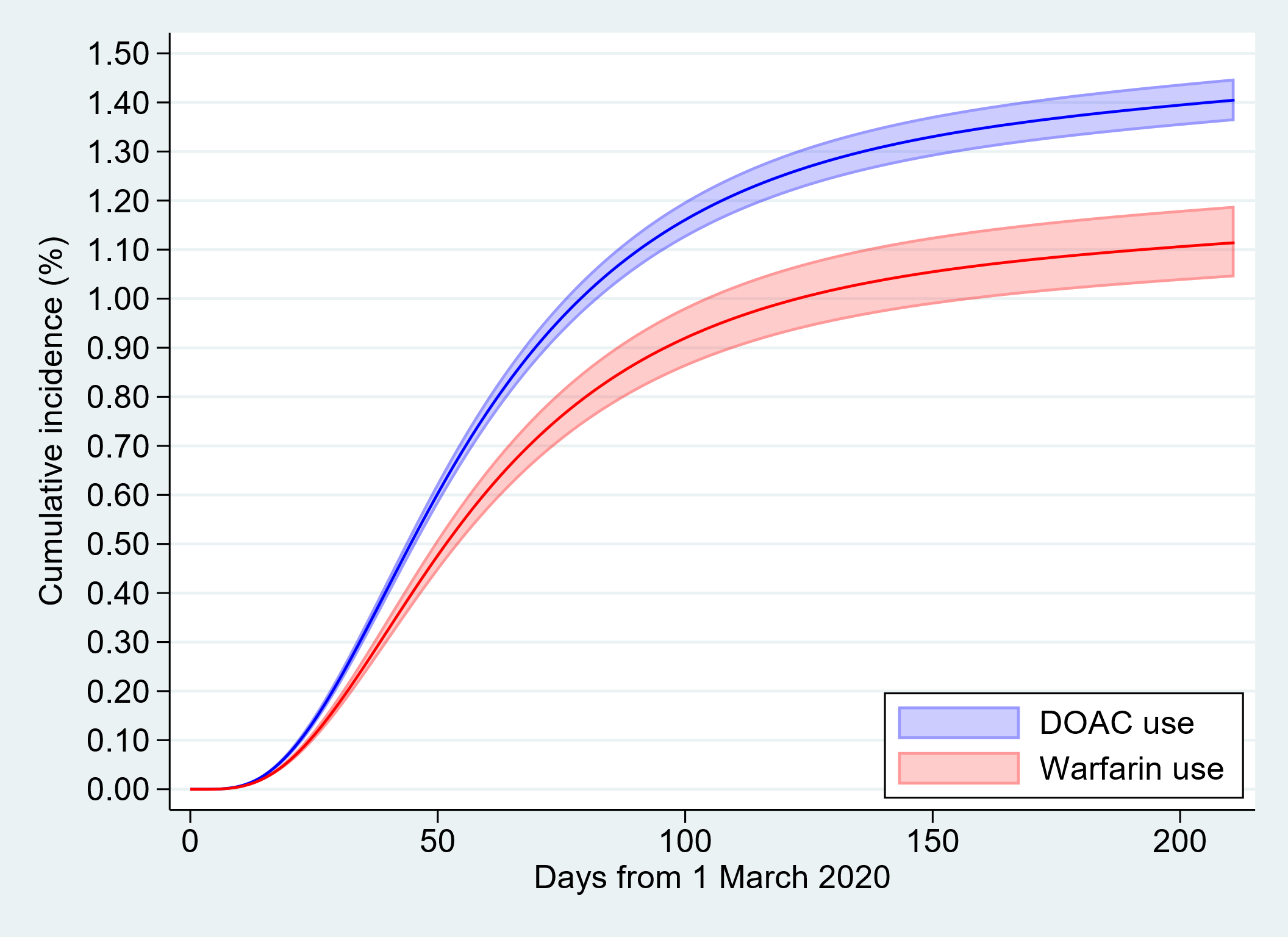
 **(b) Time to testing positive for SARS-CoV-2**

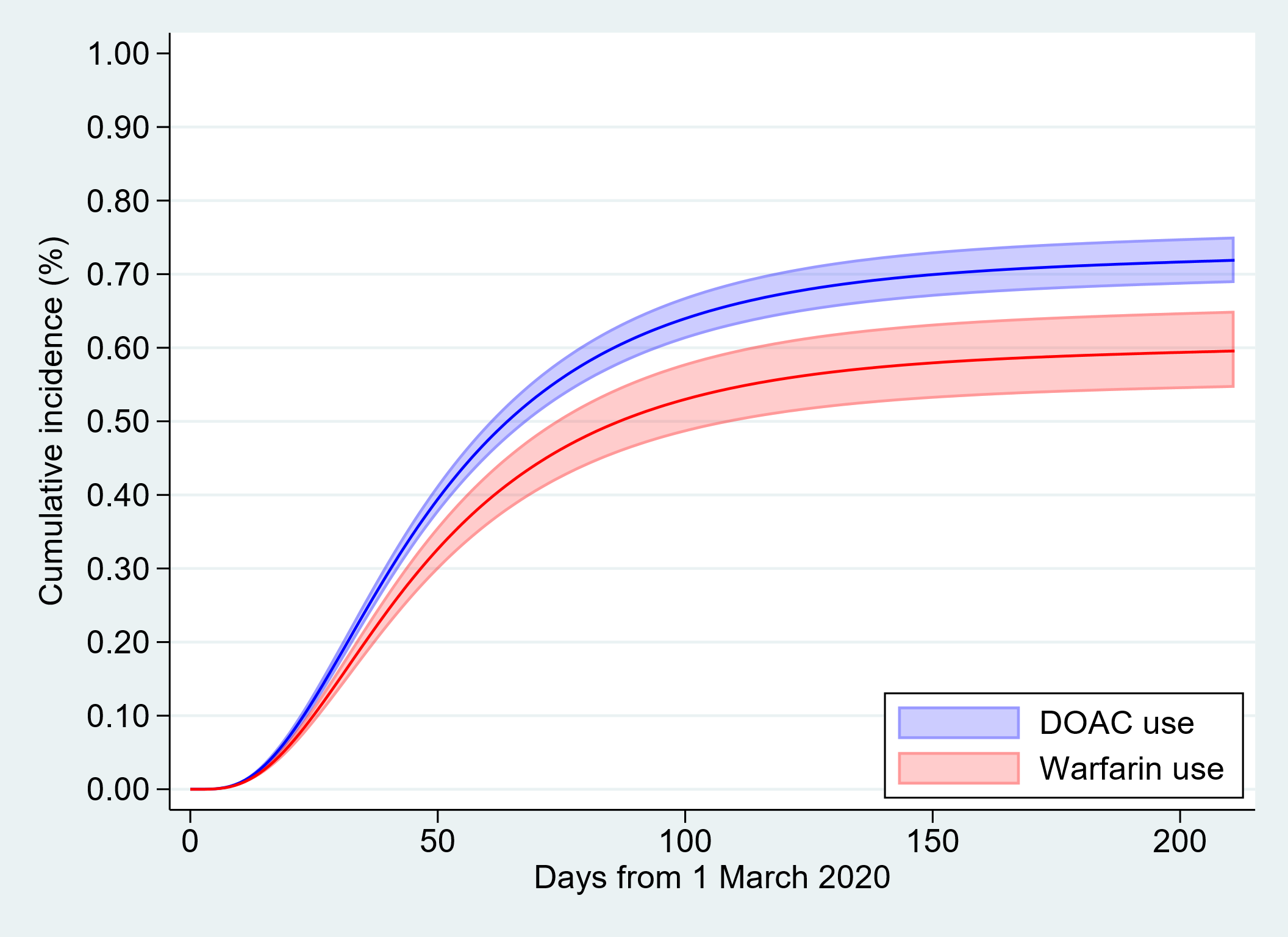
 **(c) Time to COVID-19 related hospital admission**

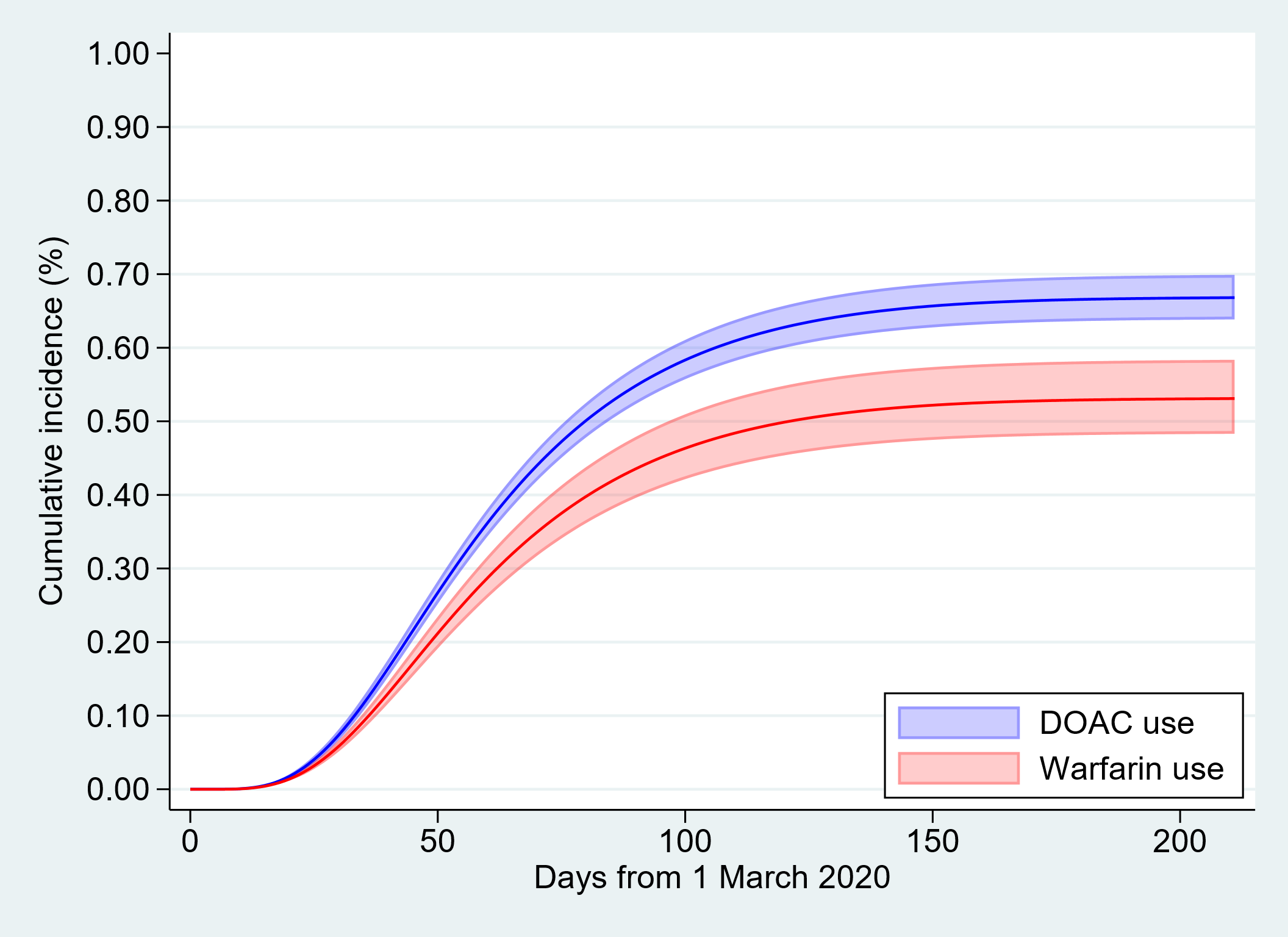
 **(d) Time to COVID-19 related deaths**

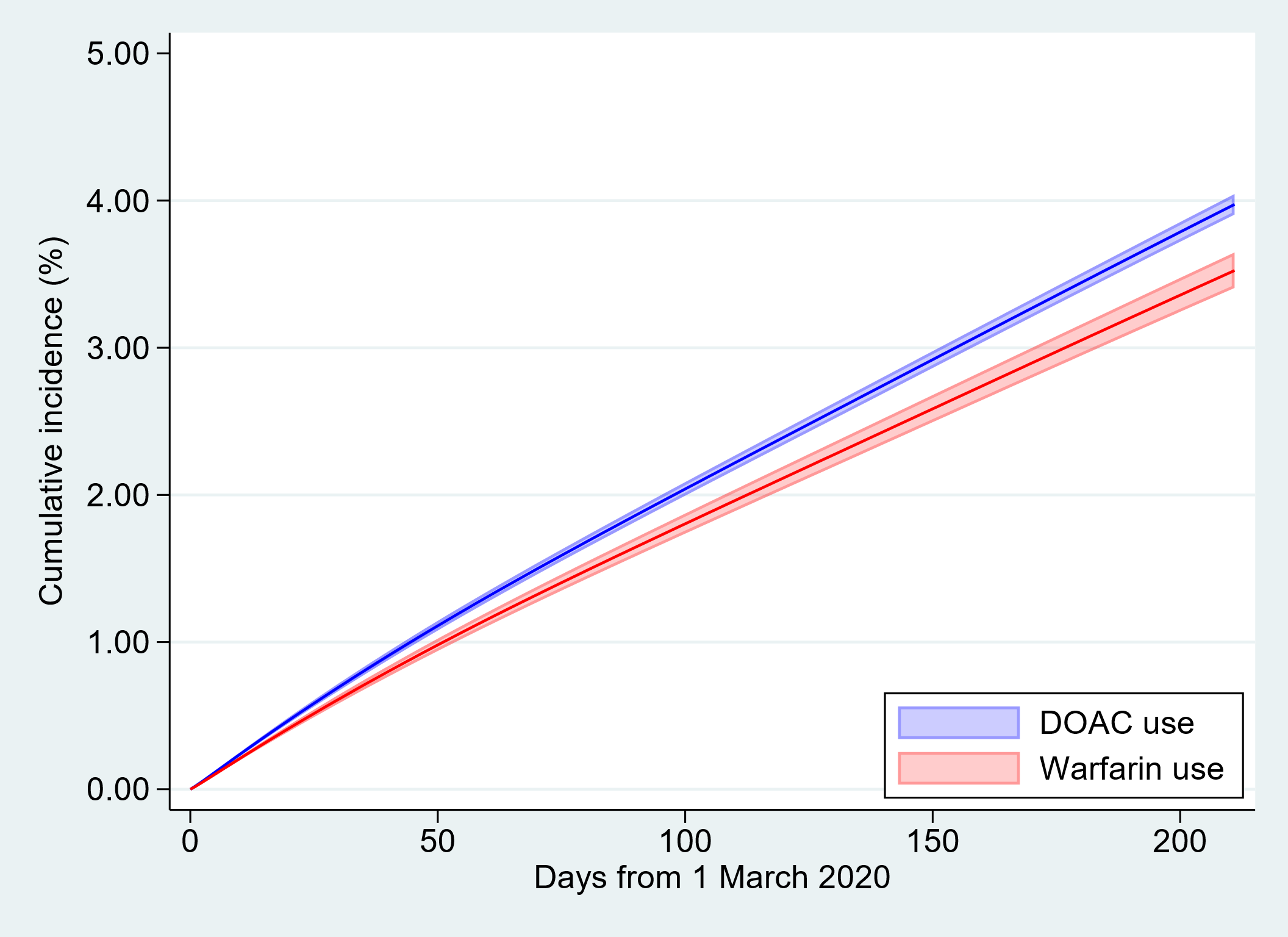
 **(e) Time to non-COVID-19 related deaths**

These figures present cumulative incidence and mortality predicted from a Royston-Parmar model including all covariates from the fully-adjusted Cox model, with the baseline hazard parametrized as a 2-degrees-of-freedom cubic spline for COVID-19 related hospital admissions, testing positive for SARS-CoV-2, and COVID-19 related deaths, 3-degrees-of-freedom cubic spline for being tested for SARS-CoV-2, and non-COVID-19 deaths; predictions standardized to the covariate distribution of the exposure group.

### **Supplementary table 3.** Results in the DAG/Fully adjusted models comparing warfarin use with direct oral anticoagulant use in people with non-valvular atrial fibrillation in Study 2.

|  | Number of events | Total person-weeks | Rate per 1,000 | Univariable | | Age/Sex Adjusted | | DAG Adjusted* | | Fully adjusted^a^ | |
| --- | --- | --- | --- | --- | --- | --- | --- | --- | --- | --- | --- |
|  |  |  |  | HR | 95% CI | HR | 95% CI | HR | 95% CI | HR | 95% CI |
| **Tested for SARS-CoV-2** | |  |  |  |  |  |  |  |  |  |  |
| DOAC use | 58123 | 7558624 | 7.69 | 1.00 (ref) |  | 1.00 (ref) |  | 1.00 (ref) |  | 1.00 (ref) |  |
| warfarin use | 15354 | 2559790 | 6.00 | 0.77 | 0.76 - 0.79 | 0.76 | 0.75 - 0.78 | 0.80 | 0.79 - 0.81 | 0.85 | 0.84 - 0.87 |
| **Testing positive for SARS-CoV-2** | |  |  |  |  |  |  |  |  |  |  |
| DOAC use | 3882 | 8148585 | 0.48 | 1.00 (ref) |  | 1.00 (ref) |  | 1.00 (ref) |  | 1.00 (ref) |  |
| warfarin use | 890 | 2709103 | 0.33 | 0.69 | 0.64 - 0.74 | 0.66 | 0.61 - 0.71 | 0.73 | 0.68 - 0.79 | 0.8 | 0.74 - 0.86 |
| **COVID-19 related hospital admission** | | |  |  |  |  |  |  |  |  |  |
| DOAC use | 1973 | 8173823 | 0.24 | 1.00 (ref) |  | 1.00 (ref) |  | 1.00 (ref) |  | 1.00 (ref) |  |
| warfarin use | 504 | 2714006 | 0.19 | 0.77 | 0.70 - 0.85 | 0.72 | 0.65 - 0.79 | 0.75 | 0.68 - 0.83 | 0.83 | 0.75 - 0.92 |
| **COVID-19 death** | |  |  |  |  |  |  |  |  |  |  |
| DOAC use | 1827 | 8195719 | 0.22 | 1.00 (ref) |  | 1.00 (ref) |  | 1.00 (ref) |  | 1.00 (ref) |  |
| warfarin use | 431 | 2719891 | 0.16 | 0.71 | 0.64 - 0.79 | 0.65 | 0.59 - 0.72 | 0.74 | 0.66 - 0.83 | 0.81 | 0.73 - 0.91 |
| **Non COVID-19 death** | |  |  |  |  |  |  |  |  |  |  |
| DOAC use | 11188 | 8195719 | 1.37 | 1.00 (ref) |  | 1.00 (ref) |  | 1.00 (ref) |  | 1.00 (ref) |  |
| warfarin use | 3007 | 2719891 | 1.11 | 0.81 | 0.78 - 0.84 | 0.76 | 0.73 - 0.79 | 0.79 | 0.76 - 0.83 | 0.88 | 0.84 - 0.92 |
| **Myocardial infarction death^b^** |  |  |  |  |  |  |  |  |  |  |  |
| DOAC use | 413 | 8195719 | 0.05 | 1.00 (ref) |  | 1.00 (ref) |  | 1.00 (ref) |  | 1.00 (ref) |  |
| warfarin use | 129 | 2719891 | 0.05 | 0.94 | 0.77 - 1.15 | 0.85 | 0.70 - 1.04 | 0.89 | 0.72 - 1.09 | 0.94 | 0.77 - 1.16 |
| **Ischemic stroke death^c,d^** |  |  |  |  |  |  |  |  |  |  |  |
| DOAC use | 64 | 8195719 | 0.01 | 1.00 (ref) |  | 1.00 (ref) |  | 1.00 (ref) |  | 1.00 (ref) |  |
| warfarin use | 11 | 2719891 | <0.01 | 0.52 | 0.27 - 0.98 | 0.48 | 0.25 - 0.91 | 0.58 | 0.30 - 1.13 | 0.63 | 0.32 - 1.22 |
| **Venous thromboembolism death^e^** |  |  |  |  |  |  |  |  |  |  |  |
| DOAC use | 21 | 8195719 | <0.01 | 1.00 (ref) |  | 1.00 (ref) |  | 1.00 (ref) |  | 1.00 (ref) |  |
| warfarin use | 6 | 2719891 | <0.01 | 0.86 | 0.35 - 2.13 | 0.83 | 0.33 - 2.06 | 0.76 | 0.30 - 1.97 | 0.80 | 0.31 - 2.07 |
| **Gastrointestinal bleed death^c^** |  |  |  |  |  |  |  |  |  |  |  |
| DOAC use | 50 | 8195719 | 0.01 | 1.00 (ref) |  | 1.00 (ref) |  | 1.00 (ref) |  | 1.00 (ref) |  |
| warfarin use | 18 | 2719891 | 0.01 | 1.08 | 0.63 - 1.86 | 1.01 | 0.59 - 1.74 | 1.07 | 0.61 - 1.88 | 1.11 | 0.63 - 1.97 |
| **Intracranial bleed death** |  |  |  |  |  |  |  |  |  |  |  |
| DOAC use | 191 | 8195719 | 0.02 | 1.00 (ref) |  | 1.00 (ref) |  | 1.00 (ref) |  | 1.00 (ref) |  |
| warfarin use | 64 | 2719891 | 0.02 | 1.01 | 0.76 - 1.34 | 0.93 | 0.70 - 1.24 | 0.97 | 0.73 - 1.31 | 1.01 | 0.75 - 1.36 |

*Adjusted for age, sex, care home residence, obesity, smoking, hypertension, heart failure, myocardial infarction, peripheral arterial disease, stroke/transient ischemic attack, venous thromboembolism, diabetes, flu vaccination, antiplatelet use, oestrogen and oestrogen-like therapy use, and Index of Multiple Deprivation and stratified on general practice.

^a^Adjusted for age, sex, care home residence, obesity, smoking, hypertension, heart failure, myocardial infarction, peripheral arterial disease, stroke/transient ischemic attack, venous thromboembolism, diabetes, flu vaccination, antiplatelet use, oestrogen and oestrogen-like therapy use, Index of Multiple Deprivation, chronic obstructive pulmonary disease, other respiratory diseases, cancer, immunosuppression, chronic kidney disease, general practice attendance and A&E attendance and stratified on general practice.

^b^Due to low event count for parameters of oestrogen use, it did not converge in the model and was dropped from the main analysis.

^c^For outcomes of gastrointestinal bleed death and ischaemic stroke death, we classified people with a diabetes diagnosis but not having HbA1c measures in the past year as uncontrolled diabetes in DAG adjusted and fully adjusted models.as the parameter for not having HbA1c measures did not converge and people with a diabetes diagnosis but not having HbA1c measures in the past year, are likely to have uncontrolled diabetes due to their potential lack of monitoring and management of diabetes.

^d^Due to low event count for parameter of immunodeficiency, it did not converge in the model and was dropped from the main analysis.

^e^Due to low event count for parameter of antiplatelet use, and immunodeficiency, they did not converge in the model and were dropped from the main analysis.

### Supplementary table 4. Additionally adjusted for ethnicity in DAG models in Study 1.

|  | **Number of events** | **Total person-weeks** | **Rate per 1,000** | **Unadjusted** | | **Age/Sex Adjusted** | | **DAG Adjusted with ethnicity*** | | **DAG Adjusted without ethnicity^a^** | | **Fully Adjusted with ethnicity^b^** | | **Fully Adjusted without ethnicity^c^** | |
| --- | --- | --- | --- | --- | --- | --- | --- | --- | --- | --- | --- | --- | --- | --- | --- |
|  |  |  |  | HR | 95% CI | HR | 95% CI | HR | 95% CI | HR | 95% CI | HR | 95% CI | HR | 95% CI |
| **Tested for SARS-CoV-2** | |  |  |  |  |  |  |  |  |  |  |  |  |  |  |
| non-use | 2202 | 375805 | 5.86 | 1.00 (ref) |  | 1.00 (ref) |  | 1.00 (ref) |  | 1.00 (ref) |  | 1.00 (ref) |  | 1.00 (ref) |  |
| current use | 6195 | 1103105 | 5.62 | 0.96 | 0.91 - 1.00 | 0.99 | 0.94 - 1.04 | 1.01 | 0.96 - 1.06 | 1.01 | 0.96 - 1.07 | 0.96 | 0.91 - 1.02 | 0.96 | 0.91 - 1.02 |
| **Testing positive for SARS-CoV-2^e^** | |  |  |  |  |  |  |  |  |  |  |  |  |  |  |
| non-use | 108 | 398879 | 0.27 | 1.00 (ref) |  | 1.00 (ref) |  | 1.00 (ref) |  | 1.00 (ref) |  | 1.00 (ref) |  | 1.00 (ref) |  |
| current use | 251 | 1165998 | 0.22 | 0.80 | 0.64 - 1.00 | 0.82 | 0.65 - 1.03 | 0.75 | 0.59 - 0.95 | 0.75 | 0.59 - 0.96 | 0.72 | 0.56 - 0.94 | 0.73 | 0.56 - 0.95 |

For outcomes of COVID-19 related hospital admission and COVID-19 death, parameters of ethnicity did not converge.

*Adjusted for age, sex, ethnicity, obesity, smoking, hypertension, heart failure, myocardial infarction, peripheral arterial disease, stroke/transient ischemic attack, venous thromboembolism, diabetes, flu vaccination, antiplatelet use, oestrogen and oestrogen-like therapy use, and Index of Multiple Deprivation.

^a^Adjusted for age, sex, obesity, smoking, hypertension, heart failure, myocardial infarction, peripheral arterial disease, stroke/transient ischemic attack, venous thromboembolism, diabetes, flu vaccination, antiplatelet use, oestrogen and oestrogen-like therapy use, and Index of Multiple Deprivation.

^b^Adjusted for age, sex, ethnicity, obesity, smoking, hypertension, heart failure, myocardial infarction, peripheral arterial disease, stroke/transient ischemic attack, venous thromboembolism, diabetes, flu vaccination, antiplatelet use, oestrogen and oestrogen-like therapy use, Index of Multiple Deprivation, chronic obstructive pulmonary disease, other respiratory diseases, cancer, immunosuppression, chronic kidney disease, general practice attendance and A&E attendance and stratified on general practice.

^c^Adjusted for age, sex, obesity, smoking, hypertension, heart failure, myocardial infarction, peripheral arterial disease, stroke/transient ischemic attack, venous thromboembolism, diabetes, flu vaccination, antiplatelet use, oestrogen and oestrogen-like therapy use, Index of Multiple Deprivation, chronic obstructive pulmonary disease, other respiratory diseases, cancer, immunosuppression, chronic kidney disease, general practice attendance and A&E attendance and stratified on general practice.

^d^We classified people with a diabetes diagnosis but not having HbA1c measures in the past year as uncontrolled diabetes in DAG adjusted and fully adjusted models.as the parameter for not having HbA1c measures did not converge and people with a diabetes diagnosis but not having HbA1c measures in the past year, are likely to have uncontrolled diabetes due to their potential lack of monitoring and management of diabetes.

### Supplementary table 5. Additionally adjusted for ethnicity in DAG models in Study 2.

|  | **Number of events** | **Total person-weeks** | **Rate per 1,000** | **Univariable** | | **Age/Sex Adjusted** | | **DAG Adjusted with ethnicity*** | | **DAG Adjusted without ethnicity^a^** | | **Fully Adjusted with ethnicity^b^** | | **Fully Adjusted without ethnicity^c^** | |
| --- | --- | --- | --- | --- | --- | --- | --- | --- | --- | --- | --- | --- | --- | --- | --- |
|  |  |  |  | HR | 95% CI | HR | 95% CI | HR | 95% CI | HR | 95% CI | HR | 95% CI | HR | 95% CI |
| **Tested for SARS-CoV-2** | |  |  |  |  |  |  |  |  |  |  |  |  |  |  |
| DOAC use | 43365 | 5604356 | 7.74 | 1.00 (ref) |  | 1.00 (ref) |  | 1.00 (ref) |  | 1.00 (ref) |  | 1.00 (ref) |  | 1.00 (ref) |  |
| warfarin use | 11367 | 1891645 | 6.01 | 0.77 | 0.76 - 0.79 | 0.76 | 0.75 - 0.78 | 0.8 | 0.78 - 0.82 | 0.8 | 0.78 - 0.82 | 0.85 | 0.84 - 0.87 | 0.85 | 0.84 - 0.87 |
| **Testing positive for SARS-CoV-2** | |  |  |  |  |  |  |  |  |  |  |  |  |  |  |
| DOAC use | 2965 | 6047824 | 0.49 | 1.00 (ref) |  | 1.00 (ref) |  | 1.00 (ref) |  | 1.00 (ref) |  | 1.00 (ref) |  | 1.00 (ref) |  |
| warfarin use | 688 | 2003268 | 0.34 | 0.70 | 0.65 - 0.76 | 0.66 | 0.61 - 0.72 | 0.74 | 0.68 - 0.81 | 0.74 | 0.68 - 0.81 | 0.81 | 0.75 - 0.89 | 0.81 | 0.75 - 0.89 |
| **COVID-19 related hospital admission** |  |  |  |  |  |  |  |  |  |  |  |  |  |  |  |
| DOAC use | 1530 | 6067451 | 0.25 | 1.00 (ref) |  | 1.00 (ref) |  | 1.00 (ref) |  | 1.00 (ref) |  | 1.00 (ref) |  | 1.00 (ref) |  |
| warfarin use | 385 | 2007241 | 0.19 | 0.76 | 0.68 - 0.85 | 0.71 | 0.63 - 0.79 | 0.75 | 0.67 - 0.84 | 0.75 | 0.67 - 0.84 | 0.83 | 0.74 - 0.94 | 0.83 | 0.74 - 0.93 |
| **COVID-19 death** | |  |  |  |  |  |  |  |  |  |  |  |  |  |  |
| DOAC use | 1345 | 6084923 | 0.22 | 1.00 (ref) |  | 1.00 (ref) |  | 1.00 (ref) |  | 1.00 (ref) |  | 1.00 (ref) |  | 1.00 (ref) |  |
| warfarin use | 318 | 2011890 | 0.16 | 0.72 | 0.63 - 0.81 | 0.65 | 0.57 - 0.73 | 0.73 | 0.64 - 0.83 | 0.73 | 0.64 - 0.83 | 0.81 | 0.71 - 0.92 | 0.81 | 0.71 - 0.92 |
| **Non COVID-19 death** | |  |  |  |  |  |  |  |  |  |  |  |  |  |  |
| DOAC use | 7800 | 6084923 | 1.28 | 1.00 (ref) |  | 1.00 (ref) |  | 1.00 (ref) |  | 1.00 (ref) |  | 1.00 (ref) |  | 1.00 (ref) |  |
| warfarin use | 2109 | 2011890 | 1.05 | 0.82 | 0.78 - 0.86 | 0.76 | 0.72 - 0.80 | 0.8 | 0.76 - 0.84 | 0.8 | 0.76 - 0.84 | 0.89 | 0.85 - 0.93 | 0.89 | 0.85 - 0.93 |

*Adjusted for age, sex, ethnicity, care home residence, obesity, smoking, hypertension, heart failure, myocardial infarction, peripheral arterial disease, stroke/transient ischemic attack, venous thromboembolism, diabetes, flu vaccination, antiplatelet use, oestrogen and oestrogen-like therapy use, and Index of Multiple Deprivation and stratified on general practice.

^a^Adjusted for age, sex, care home residence, obesity, smoking, hypertension, heart failure, myocardial infarction, peripheral arterial disease, stroke/transient ischemic attack, venous thromboembolism, diabetes, flu vaccination, antiplatelet use, oestrogen and oestrogen-like therapy use, and Index of Multiple Deprivation and stratified on general practice

^b^Adjusted for age, sex, ethnicity, care home residence, obesity, smoking, hypertension, heart failure, myocardial infarction, peripheral arterial disease, stroke/transient ischemic attack, venous thromboembolism, diabetes, flu vaccination, antiplatelet use, oestrogen and oestrogen-like therapy use, Index of Multiple Deprivation, chronic obstructive pulmonary disease, other respiratory diseases, cancer, immunosuppression, chronic kidney disease, general practice attendance and A&E attendance and stratified on general practice.

^c^Adjusted for age, sex, care home residence, obesity, smoking, hypertension, heart failure, myocardial infarction, peripheral arterial disease, stroke/transient ischemic attack, venous thromboembolism, diabetes, flu vaccination, antiplatelet use, oestrogen and oestrogen-like therapy use, Index of Multiple Deprivation, chronic obstructive pulmonary disease, other respiratory diseases, cancer, immunosuppression, chronic kidney disease, general practice attendance and A&E attendance and stratified on general practice.

### Supplementary table 6. Excluded people prescribed antiplatelets 4 months before study start date in Study 1.

|  | **Number of events** | **Total person-weeks** | **Rate per 1,000** | **Unadjusted** | | **Age/Sex Adjusted** | | **DAG Adjusted*** | | **Fully adjusted^a^** | | |
| --- | --- | --- | --- | --- | --- | --- | --- | --- | --- | --- | --- | --- |
|  |  |  |  | HR | 95% CI | HR | 95% CI | HR | 95% CI | | HR | 95% CI |
| **Tested for SARS-CoV-2** |  |  |  |  |  |  |  |  |  | |  |  |
| non-use | 2292 | 390116 | 5.88 | 1.00 (ref) |  | 1.00 (ref) |  | 1.00 (ref) |  | | 1.00 (ref) |  |
| current use | 7779 | 1410757 | 5.51 | 0.93 | 0.89 - 0.98 | 0.97 | 0.93 - 1.02 | 0.96 | 0.92 - 1.01 | | 0.93 | 0.88 - 0.98 |
| **Testing positive for SARS-CoV-2** | |  |  |  |  |  |  |  |  | |  |  |
| non-use | 119 | 414366 | 0.29 | 1.00 (ref) |  | 1.00 (ref) |  | 1.00 (ref) |  | | 1.00 (ref) |  |
| current use | 323 | 1488272 | 0.22 | 0.76 | 0.61 - 0.93 | 0.78 | 0.63 - 0.97 | 0.77 | 0.62 - 0.95 | | 0.73 | 0.58 - 0.92 |
| **COVID-19 related hospital admission^b^** |  |  |  |  |  |  |  |  |  | |  |  |
| non-use | 46 | 415326 | 0.11 | 1.00 (ref) |  | 1.00 (ref) |  | 1.00 (ref) |  | | 1.00 (ref) |  |
| current use | 163 | 1490392 | 0.11 | 0.99 | 0.71 - 1.37 | 0.95 | 0.68 - 1.33 | 0.91 | 0.65 - 1.27 | | 0.84 | 0.59 - 1.21 |
| **COVID-19 death^b,c^** | |  |  |  |  |  |  |  |  | |  |  |
| non-use | 44 | 415960 | 0.11 | 1.00 (ref) |  | 1.00 (ref) |  | 1.00 (ref) |  | | 1.00 (ref) |  |
| current use | 128 | 1492404 | 0.09 | 0.81 | 0.58 - 1.15 | 0.77 | 0.55 - 1.09 | 0.72 | 0.51 - 1.03 | | 0.74 | 0.50 - 1.11 |

*Adjusted for age, sex, obesity, smoking, hypertension, heart failure, myocardial infarction, peripheral arterial disease, stroke/transient ischemic attack, venous thromboembolism, diabetes, flu vaccination, oestrogen and oestrogen-like therapy use, and Index of Multiple Deprivation.

^a^Adjusted for age, sex, obesity, smoking, hypertension, heart failure, myocardial infarction, peripheral arterial disease, stroke/transient ischemic attack, venous thromboembolism, diabetes, flu vaccination, oestrogen and oestrogen-like therapy use, Index of Multiple Deprivation, chronic obstructive pulmonary disease, other respiratory diseases, cancer, immunosuppression, chronic kidney disease, general practice attendance and A&E attendance and stratified on general practice.

^b^For outcomes of COVID-19 related hospital admission and COVID-19 death, we classified people with a diabetes diagnosis but not having HbA1c measures in the past year as uncontrolled diabetes in DAG adjusted and fully adjusted models.as the parameter for not having HbA1c measures did not converge and people with a diabetes diagnosis but not having HbA1c measures in the past year, are likely to have uncontrolled diabetes due to their potential lack of monitoring and management of diabetes.

^c^Due to low event count for parameters of stroke/transient ischaemic attack, oestrogen use and peripheral artery disease, they did not converge in the model and were dropped from the main analysis.

### Supplementary table 7. Excluded people prescribed antiplatelets 4 months before study start date in Study 2.

|  | **Number of events** | **Total person-weeks** | **Rate per 1,000** | **Unadjusted** | | **Age/Sex Adjusted** | | **DAG Adjusted*** | | **Fully adjusted^a^** | |
| --- | --- | --- | --- | --- | --- | --- | --- | --- | --- | --- | --- |
|  |  |  |  | HR | 95% CI | HR | 95% CI | HR | 95% CI | HR | 95% CI |
| **Tested for SARS-CoV-2** |  |  |  |  |  |  |  |  |  |  |  |
| non-use | 53304 | 7057738 | 7.55 | 1.00 (ref) |  | 1.00 (ref) |  | 1.00 (ref) |  | 1.00 (ref) |  |
| current use | 14451 | 2449231 | 5.9 | 0.78 | 0.76 - 0.79 | 0.77 | 0.75 - 0.78 | 0.8 | 0.78 - 0.81 | 0.85 | 0.83 - 0.87 |
| **Testing positive for**  **SARS-CoV-2** | |  |  |  |  |  |  |  |  |  |  |
| non-use | 3598 | 7597766 | 0.47 | 1.00 (ref) |  | 1.00 (ref) |  | 1.00 (ref) |  | 1.00 (ref) |  |
| current use | 852 | 2589587 | 0.33 | 0.7 | 0.65 - 0.75 | 0.66 | 0.61 - 0.71 | 0.74 | 0.68 - 0.80 | 0.8 | 0.74 - 0.87 |
| **COVID-19 related hospital admission** |  |  |  |  |  |  |  |  |  |  |  |
| non-use | 1793 | 7621589 | 0.24 | 1.00 (ref) |  | 1.00 (ref) |  | 1.00 (ref) |  | 1.00 (ref) |  |
| current use | 477 | 2594380 | 0.18 | 0.78 | 0.71 - 0.87 | 0.73 | 0.66 - 0.80 | 0.75 | 0.68 - 0.84 | 0.83 | 0.75 - 0.92 |
| **COVID-19 death** | |  |  |  |  |  |  |  |  |  |  |
| non-use | 1700 | 7641320 | 0.22 | 1.00 (ref) |  | 1.00 (ref) |  | 1.00 (ref) |  | 1.00 (ref) |  |
| current use | 410 | 2599887 | 0.16 | 0.71 | 0.64 - 0.79 | 0.65 | 0.58 - 0.72 | 0.73 | 0.66 - 0.82 | 0.8 | 0.72 - 0.90 |
| **Non-COVID-19 death** |  |  |  |  |  |  |  |  |  |  |  |
| non-use | 10331 | 7641320 | 1.35 | 1.00 (ref) |  | 1.00 (ref) |  | 1.00 (ref) |  | 1.00 (ref) |  |
| current use | 2821 | 2599887 | 1.09 | 0.8 | 0.77 - 0.84 | 0.75 | 0.72 - 0.78 | 0.78 | 0.75 - 0.82 | 0.87 | 0.83 - 0.90 |

*Adjusted for age, sex, care home residence, obesity, smoking, hypertension, heart failure, myocardial infarction, peripheral arterial disease, stroke/transient ischemic attack, venous thromboembolism, diabetes, flu vaccination, oestrogen and oestrogen-like therapy use, and Index of Multiple Deprivation and stratified on general practice.

^a^Adjusted for age, sex, care home residence, obesity, smoking, hypertension, heart failure, myocardial infarction, peripheral arterial disease, stroke/transient ischemic attack, venous thromboembolism, diabetes, flu vaccination, oestrogen and oestrogen-like therapy use, Index of Multiple Deprivation, chronic obstructive pulmonary disease, other respiratory diseases, cancer, immunosuppression, chronic kidney disease, general practice attendance and A&E attendance and stratified on general practice.

### Supplementary table 8. Limited the study cohort to people who aged 55 or above in Study 1.

|  | **Number of events** | **Total person-weeks** | **Rate per 1,000** | **Unadjusted** | | **Age/Sex Adjusted** | | **DAG Adjusted*** | | **Fully adjusted^a^** | | |
| --- | --- | --- | --- | --- | --- | --- | --- | --- | --- | --- | --- | --- |
|  |  |  |  | HR | 95% CI | HR | 95% CI | HR | 95% CI | | HR | 95% CI |
| **Tested for SARS-CoV-2** |  |  |  |  |  |  |  |  |  | |  |  |
| non-use | 2593 | 455564 | 5.69 | 1.00 (ref) |  | 1.00 (ref) |  | 1.00 (ref) |  | | 1.00 (ref) |  |
| current use | 7813 | 1408949 | 5.55 | 0.97 | 0.93 - 1.02 | 0.99 | 0.95 - 1.04 | 1.01 | 0.97 - 1.06 | | 0.98 | 0.93 - 1.03 |
| **Testing positive for SARS-CoV-2** | |  |  |  |  |  |  |  |  | |  |  |
| non-use | 139 | 482272 | 0.29 | 1.00 (ref) |  | 1.00 (ref) |  | 1.00 (ref) |  | | 1.00 (ref) |  |
| current use | 315 | 1486786 | 0.21 | 0.74 | 0.60 - 0.90 | 0.77 | 0.63 - 0.94 | 0.73 | 0.59 - 0.90 | | 0.69 | 0.55 - 0.87 |
| **COVID-19 related hospital admission ^b^** |  |  |  |  |  |  |  |  |  | |  |  |
| non-use | 58 | 483349 | 0.12 | 1.00 (ref) |  | 1.00 (ref) |  | 1.00 (ref) |  | | 1.00 (ref) |  |
| current use | 161 | 1488862 | 0.11 | 0.90 | 0.67 - 1.22 | 0.90 | 0.67 - 1.22 | 0.85 | 0.61 - 1.17 | | 0.78 | 0.55 - 1.09 |
| **COVID-19 death^b,c^** | |  |  |  |  |  |  |  |  | |  |  |
| non-use | 54 | 484185 | 0.11 | 1.00 (ref) |  | 1.00 (ref) |  | 1.00 (ref) |  | | 1.00 (ref) |  |
| current use | 129 | 1490798 | 0.09 | 0.78 | 0.57 - 1.07 | 0.78 | 0.57 - 1.08 | 0.70 | 0.50 - 0.97 | | 0.67 | 0.46 - 0.98 |

*Adjusted for age, sex, obesity, smoking, hypertension, heart failure, myocardial infarction, peripheral arterial disease, stroke/transient ischemic attack, venous thromboembolism, diabetes, flu vaccination, antiplatelet use, oestrogen and oestrogen-like therapy use, and Index of Multiple Deprivation.

^a^Adjusted for age, sex, obesity, smoking, hypertension, heart failure, myocardial infarction, peripheral arterial disease, stroke/transient ischemic attack, venous thromboembolism, diabetes, flu vaccination, antiplatelet use, oestrogen and oestrogen-like therapy use, Index of Multiple Deprivation, chronic obstructive pulmonary disease, other respiratory diseases, cancer, immunosuppression, chronic kidney disease, general practice attendance and A&E attendance and stratified on general practice.

^b^For outcomes of COVID-19 related hospital admission and COVID-19 death, we classified people with a diabetes diagnosis but not having HbA1c measures in the past year as uncontrolled diabetes in DAG adjusted and fully adjusted models.as the parameter for not having HbA1c measures did not converge and people with a diabetes diagnosis but not having HbA1c measures in the past year, are likely to have uncontrolled diabetes due to their potential lack of monitoring and management of diabetes.

^c^Due to low event count for parameters of stroke/transient ischaemic attack, and oestrogen use, they did not converge in the model and were dropped from the main analysis.

### Supplementary table 9. Subgroup analysis on positive COVID-test according to care home residence in the DAG and fully adjusted models comparing current use with non-user in Study 1.

|  | **Number of events** | **Total person-weeks** | **Rate per 1,000** | **Unadjusted** | | **Age/Sex Adjusted** | | **DAG Adjusted*** | | **Fully adjusted^a^** | | |
| --- | --- | --- | --- | --- | --- | --- | --- | --- | --- | --- | --- | --- |
|  |  |  |  | HR | 95% CI | HR | 95% CI | HR | 95% CI | | HR | 95% CI |
|  |  |  |  |  | P(interaction)= 0.457 |  | P(interaction)= 0.315 |  | P(interaction)= 0.332 | |  | P(interaction)= 0.804 |
| **Not living in care-home** | |  |  |  |  |  |  |  |  | |  |  |
| non-use | 131 | 526491 | 0.25 | 1.00 (ref) | | 1.00 (ref) | | 1.00 (ref) | | | 1.00 (ref) | |
| current use | 301 | 1548828 | 0.19 | 0.78 | 0.64 - 0.96 | 0.80 | 0.65 - 0.98 | 0.75 | 0.60 - 0.93 | | 0.70 | 0.55 - 0.87 |
| **Living in care-home** | |  |  |  |  |  |  |  |  | |  |  |
| non-use | 19 | 4815 | 3.95 | 1.00 (ref) | | 1.00 (ref) | | 1.00 (ref) | | | 1.00 (ref) | |
| current use | 30 | 7854 | 3.82 | 0.98 | 0.55 - 1.75 | 1.09 | 0.61 - 1.94 | 1.01 | 0.57 - 1.80 | | 0.77 | 0.36 - 1.65 |

*Adjusted for age, sex, obesity, smoking, hypertension, heart failure, myocardial infarction, peripheral arterial disease, stroke/transient ischemic attack, venous thromboembolism, diabetes, flu vaccination, antiplatelet use, oestrogen and oestrogen-like therapy use, and Index of Multiple Deprivation.

^a^Adjusted for age, sex, obesity, smoking, hypertension, heart failure, myocardial infarction, peripheral arterial disease, stroke/transient ischemic attack, venous thromboembolism, diabetes, flu vaccination, antiplatelet use, oestrogen and oestrogen-like therapy use, Index of Multiple Deprivation, chronic obstructive pulmonary disease, other respiratory diseases, cancer, immunosuppression, chronic kidney disease, general practice attendance and A&E attendance and stratified on general practice.

### Supplementary table 10. Excluded people who were prescribed both warfarin and direct oral anticoagulants on the day as the latest oral anticoagulant prescription in Study 2.

|  | **Number of events** | **Total person-weeks** | **Rate per 1,000** | **Unadjusted** | | **Age/Sex Adjusted** | | **DAG Adjusted*** | | **Fully adjusted^a^** | | |
| --- | --- | --- | --- | --- | --- | --- | --- | --- | --- | --- | --- | --- |
|  |  |  |  | HR | 95% CI | HR | 95% CI | HR | 95% CI | | HR | 95% CI |
| **Tested for SARS-CoV-2** |  |  |  |  |  |  |  |  |  | |  |  |
| non-use | 58123 | 7558624 | 7.69 | 1.00 (ref) |  | 1.00 (ref) |  | 1.00 (ref) |  | | 1.00 (ref) |  |
| current use | 15346 | 2558921 | 6.00 | 0.77 | 0.76 - 0.79 | 0.76 | 0.75 - 0.78 | 0.8 | 0.79 - 0.81 | | 0.85 | 0.84 - 0.87 |
| **Testing positive for SARS-CoV-2** | |  |  |  |  |  |  |  |  | |  |  |
| non-use | 3882 | 8148585 | 0.48 | 1.00 (ref) |  | 1.00 (ref) |  | 1.00 (ref) |  | | 1.00 (ref) |  |
| current use | 889 | 2708170 | 0.33 | 0.69 | 0.64 - 0.74 | 0.66 | 0.61 - 0.71 | 0.73 | 0.68 - 0.79 | | 0.8 | 0.74 - 0.86 |
| **COVID-19 related hospital admission** |  |  |  |  |  |  |  |  |  | |  |  |
| non-use | 1973 | 8173823 | 0.24 | 1.00 (ref) |  | 1.00 (ref) |  | 1.00 (ref) |  | | 1.00 (ref) |  |
| current use | 504 | 2713072 | 0.19 | 0.77 | 0.70 - 0.85 | 0.72 | 0.65 - 0.79 | 0.75 | 0.68 - 0.83 | | 0.83 | 0.75 - 0.92 |
| **COVID-19 death** | |  |  |  |  |  |  |  |  | |  |  |
| non-use | 1827 | 8195719 | 0.22 | 1.00 (ref) |  | 1.00 (ref) |  | 1.00 (ref) |  | | 1.00 (ref) |  |
| current use | 430 | 2718957 | 0.16 | 0.71 | 0.64 - 0.79 | 0.65 | 0.58 - 0.72 | 0.74 | 0.66 - 0.82 | | 0.81 | 0.72 - 0.90 |
| **Non-COVID-19 death** |  |  |  |  |  |  |  |  |  | |  |  |
| non-use | 11188 | 8195719 | 1.37 | 1.00 (ref) |  | 1.00 (ref) |  | 1.00 (ref) |  | | 1.00 (ref) |  |
| current use | 3005 | 2718957 | 1.11 | 0.81 | 0.78 - 0.84 | 0.76 | 0.73 - 0.79 | 0.79 | 0.76 - 0.83 | | 0.88 | 0.84 - 0.92 |

*Adjusted for age, sex, care home residence, obesity, smoking, hypertension, heart failure, myocardial infarction, peripheral arterial disease, stroke/transient ischemic attack, venous thromboembolism, diabetes, flu vaccination, antiplatelet use, oestrogen and oestrogen-like therapy use, and Index of Multiple Deprivation and stratified on general practice.

^a^Adjusted for age, sex, care home residence, obesity, smoking, hypertension, heart failure, myocardial infarction, peripheral arterial disease, stroke/transient ischemic attack, venous thromboembolism, diabetes, flu vaccination, antiplatelet use, oestrogen and oestrogen-like therapy use, Index of Multiple Deprivation, chronic obstructive pulmonary disease, other respiratory diseases, cancer, immunosuppression, chronic kidney disease, general practice attendance and A&E attendance and stratified on general practice.

### Supplementary table 11. Excluded people who ever had warfarin prescription 4 months before study start date in the direct oral anticoagulant group in Study 2.

|  | **Number of events** | **Total person-weeks** | **Rate per 1,000** | **Unadjusted** | | **Age/Sex Adjusted** | | **DAG Adjusted*** | | **Fully adjusted^a^** | | |
| --- | --- | --- | --- | --- | --- | --- | --- | --- | --- | --- | --- | --- |
|  |  |  |  | HR | 95% CI | HR | 95% CI | HR | 95% CI | | HR | 95% CI |
| **Tested for SARS-CoV-2** |  |  |  |  |  |  |  |  |  | |  |  |
| non-use | 57285 | 7469075 | 7.67 | 1.00 (ref) |  | 1.00 (ref) |  | 1.00 (ref) |  | | 1.00 (ref) |  |
| current use | 15354 | 2559790 | 6 | 0.78 | 0.76 - 0.79 | 0.77 | 0.75 - 0.78 | 0.8 | 0.79 - 0.82 | | 0.86 | 0.84 - 0.87 |
| **Testing positive for SARS-CoV-2** | |  |  |  |  |  |  |  |  | |  |  |
| non-use | 3804 | 8050487 | 0.47 | 1.00 (ref) |  | 1.00 (ref) |  | 1.00 (ref) |  | | 1.00 (ref) |  |
| current use | 890 | 2709103 | 0.33 | 0.7 | 0.65 - 0.75 | 0.66 | 0.62 - 0.71 | 0.74 | 0.69 - 0.80 | | 0.81 | 0.75 - 0.87 |
| **COVID-19 related hospital admission** |  |  |  |  |  |  |  |  |  | |  |  |
| non-use | 1935 | 8075203 | 0.24 | 1.00 (ref) |  | 1.00 (ref) |  | 1.00 (ref) |  | | 1.00 (ref) |  |
| current use | 504 | 2714006 | 0.19 | 0.78 | 0.70 - 0.86 | 0.72 | 0.65 - 0.80 | 0.76 | 0.68 - 0.84 | | 0.84 | 0.76 - 0.93 |
| **COVID-19 death** | |  |  |  |  |  |  |  |  | |  |  |
| non-use | 1792 | 8096758 | 0.22 | 1.00 (ref) |  | 1.00 (ref) |  | 1.00 (ref) |  | | 1.00 (ref) |  |
| current use | 431 | 2719891 | 0.16 | 0.72 | 0.65 - 0.80 | 0.65 | 0.59 - 0.73 | 0.74 | 0.67 - 0.83 | | 0.81 | 0.73 - 0.91 |
| **Non-COVID-19 death** |  |  |  |  |  |  |  |  |  | |  |  |
| non-use | 10949 | 8096758 | 1.35 | 1.00 (ref) |  | 1.00 (ref) |  | 1.00 (ref) |  | | 1.00 (ref) |  |
| current use | 3007 | 2719891 | 1.11 | 0.82 | 0.79 - 0.85 | 0.76 | 0.73 - 0.80 | 0.8 | 0.77 - 0.83 | | 0.89 | 0.85 - 0.92 |

*Adjusted for age, sex, care home residence, obesity, smoking, hypertension, heart failure, myocardial infarction, peripheral arterial disease, stroke/transient ischemic attack, venous thromboembolism, diabetes, flu vaccination, antiplatelet use, oestrogen and oestrogen-like therapy use, and Index of Multiple Deprivation and stratified on general practice.

^a^Adjusted for age, sex, care home residence, obesity, smoking, hypertension, heart failure, myocardial infarction, peripheral arterial disease, stroke/transient ischemic attack, venous thromboembolism, diabetes, flu vaccination, antiplatelet use, oestrogen and oestrogen-like therapy use, Index of Multiple Deprivation, chronic obstructive pulmonary disease, other respiratory diseases, cancer, immunosuppression, chronic kidney disease, general practice attendance and A&E attendance and stratified on general practice.

### Supplementary table 12. Time-updated oral anticoagulant exposure variable in Study 2.

|  | **Number of events** | **Total person-weeks** | **Rate per 1,000** | **Unadjusted** | | **Age/Sex Adjusted** | | **DAG Adjusted*** | | **Fully adjusted^a^** | | |
| --- | --- | --- | --- | --- | --- | --- | --- | --- | --- | --- | --- | --- |
|  |  |  |  | HR | 95% CI | HR | 95% CI | HR | 95% CI | | HR | 95% CI |
| **Tested for SARS-CoV-2** |  |  |  |  |  |  |  |  |  | |  |  |
| non-use | 60952 | 7903763 | 7.71 | 1.00 (ref) |  | 1.00 (ref) |  | 1.00 (ref) |  | | 1.00 (ref) |  |
| current use | 12525 | 2214651 | 5.66 | 0.76 | 0.74 - 0.77 | 0.75 | 0.73 - 0.76 | 0.78 | 0.76 - 0.80 | | 0.83 | 0.82 - 0.85 |
| **Testing positive for SARS-CoV-2** | |  |  |  |  |  |  |  |  | |  |  |
| non-use | 3994 | 8535512 | 0.47 | 1.00 (ref) |  | 1.00 (ref) |  | 1.00 (ref) |  | | 1.00 (ref) |  |
| current use | 778 | 2322177 | 0.34 | 0.68 | 0.63 - 0.73 | 0.65 | 0.60 - 0.70 | 0.71 | 0.66 - 0.77 | | 0.78 | 0.72 - 0.84 |
| **COVID-19 related hospital admission** |  |  |  |  |  |  |  |  |  | |  |  |
| non-use | 2025 | 8562354 | 0.24 | 1.00 (ref) |  | 1.00 (ref) |  | 1.00 (ref) |  | | 1.00 (ref) |  |
| current use | 452 | 2325475 | 0.19 | 0.76 | 0.69 - 0.84 | 0.71 | 0.64 - 0.79 | 0.73 | 0.66 - 0.81 | | 0.81 | 0.73 - 0.90 |
| **COVID-19 death** | |  |  |  |  |  |  |  |  | |  |  |
| non-use | 1888 | 8586646 | 0.22 | 1.00 (ref) |  | 1.00 (ref) |  | 1.00 (ref) |  | | 1.00 (ref) |  |
| current use | 370 | 2328963 | 0.16 | 0.69 | 0.61 - 0.77 | 0.63 | 0.57 - 0.71 | 0.7 | 0.62 - 0.79 | | 0.77 | 0.69 - 0.87 |
| **Non-COVID-19 death** |  |  |  |  |  |  |  |  |  | |  |  |
| non-use | 11802 | 8586646 | 1.37 | 1.00 (ref) |  | 1.00 (ref) |  | 1.00 (ref) |  | | 1.00 (ref) |  |
| current use | 2393 | 2328963 | 1.03 | 0.74 | 0.71 - 0.77 | 0.7 | 0.67 - 0.73 | 0.73 | 0.70 - 0.76 | | 0.81 | 0.77 - 0.85 |

*Adjusted for age, sex, care home residence, obesity, smoking, hypertension, heart failure, myocardial infarction, peripheral arterial disease, stroke/transient ischemic attack, venous thromboembolism, diabetes, flu vaccination, antiplatelet use, oestrogen and oestrogen-like therapy use, and Index of Multiple Deprivation and stratified on general practice.

^a^Adjusted for age, sex, care home residence, obesity, smoking, hypertension, heart failure, myocardial infarction, peripheral arterial disease, stroke/transient ischemic attack, venous thromboembolism, diabetes, flu vaccination, antiplatelet use, oestrogen and oestrogen-like therapy use, Index of Multiple Deprivation, chronic obstructive pulmonary disease, other respiratory diseases, cancer, immunosuppression, chronic kidney disease, general practice attendance and A&E attendance and stratified on general practice.

### Supplementary table 13. Bias-analyses for DAG-adjusted hazard ratios in Study 1.

|  | Bias-adjusted hazard ratio | Observed hazard ratio | | | E-value | | | Cornfield condition | | |
| --- | --- | --- | --- | --- | --- | --- | --- | --- | --- | --- |
|  |  | Point estimate | Lower CI bound | Upper CI bound | Point estimate | Lower CI bound | Upper CI bound | Point estimate | Lower CI bound | Upper CI bound |
| Testing positive for SARS-CoV-2 | 1 | 0.73 | 0.60 | 0.90 | 2.08 | 2.72 | 1.46 | 1.37 | 1.67 | 1.11 |
| COVID-19 related hospital admission | 1 | 0.86 | 0.63 | 1.17 | 1.60 | 2.55 | - | 1.16 | 1.59 | - |
| COVID-19 death | 1 | 0.69 | 0.49 | 0.97 | 2.26 | 3.50 | 1.21 | 1.45 | 2.04 | 1.03 |

As a simplification, we consider higher-risk health behaviour to be a binary variable and assume no interaction between the unmeasured confounder and measured covariates on the outcome. To apply the bias analysis formulas to hazard ratios, we assume the outcome is rare.

### Supplementary table 14. Bias-analyses for DAG-adjusted hazard ratios in Study 2.

|  | Bias-adjusted hazard ratio | Observed hazard ratio | | | E-value | | | Cornfield condition | | |
| --- | --- | --- | --- | --- | --- | --- | --- | --- | --- | --- |
|  |  | Point estimate | Lower CI bound | Upper CI bound | Point estimate | Lower CI bound | Upper CI bound | Point estimate | Lower CI bound | Upper CI bound |
| Tested for SARS-CoV-2 | 1 | 0.80 | 0.79 | 0.81 | 1.81 | 1.85 | 1.77 | 1.25 | 1.27 | 1.23 |
| Testing positive for SARS-CoV-2 | 1 | 0.73 | 0.68 | 0.79 | 2.08 | 2.30 | 1.85 | 1.37 | 1.47 | 1.27 |
| COVID-19 related hospital admission | 1 | 0.75 | 0.68 | 0.83 | 2.00 | 2.30 | 1.70 | 1.33 | 1.47 | 1.20 |
| COVID-19 death | 1 | 0.74 | 0.66 | 0.83 | 2.04 | 2.40 | 1.70 | 1.35 | 1.52 | 1.20 |
| Tested for SARS-CoV-2 | 1.2 | 0.80 | 0.79 | 0.81 | 2.37 | 2.41 | 2.33 | 1.50 | 1.52 | 1.48 |
| Testing positive for SARS-CoV-2 | 1.2 | 0.73 | 0.68 | 0.79 | 2.67 | 2.93 | 2.41 | 1.64 | 1.76 | 1.52 |
| COVID-19 related hospital admission | 1.2 | 0.75 | 0.68 | 0.83 | 2.58 | 2.93 | 2.25 | 1.60 | 1.76 | 1.45 |
| COVID-19 death | 1.2 | 0.74 | 0.66 | 0.83 | 2.63 | 3.04 | 2.25 | 1.62 | 1.82 | 1.45 |

As a simplification, we consider higher-risk health behaviour to be a binary variable and assume no interaction between the unmeasured confounder and measured covariates on the outcome. To apply the bias analysis formulas to hazard ratios, we assume the outcome is rare.
